# Machine learning to predict antibiotic susceptibility in Enterobacterales bloodstream infections compared to clinician prescribing

**DOI:** 10.1101/2024.10.02.24314776

**Authors:** Kevin Yuan, Augustine Luk, Jia Wei, A Sarah Walker, Tingting Zhu, David W Eyre

**Affiliations:** Big Data Institute, Nuffield Department of Population Health, University of Oxford, Oxford, UK; Nuffield Department of Medicine, University of Oxford, Oxford, UK; NIHR Health Protection Research Unit in Healthcare Associated Infections and Antimicrobial Resistance, University of Oxford, Oxford, UK; NIHR Oxford Biomedical Research Centre, Oxford, UK; Institute of Biomedical Engineering, University of Oxford, Oxford, UK; Oxford University Hospitals NHS Foundation Trust, Oxford, UK

## Abstract

**Background:** Patients with Gram-negative bloodstream infections are at risk of serious adverse outcomes without active treatment, but identifying who has antimicrobial resistance (AMR) to target empirical treatment is challenging.

**Methods:** We used XGBoost machine learning models to predict the presence of antimicrobial resistance to seven antibiotics in patients with Enterobacterales bloodstream infection. Models were trained using hospital and community data available at the time blood cultures were obtained from Oxfordshire, UK, between 01-January-2017 and 31-December-2021. Model performance was compared to final microbiology results using test datasets from 01-January-2022 to 31-December-2023 and with clinicians’ prescribing.

**Findings:** 4709 infection episodes were used for model training and evaluation; antibiotic resistance rates ranged from 7-67%. In held-out test data, resistance prediction performance was similar for the seven antibiotics (AUCs 0.680 [95%CI 0.641-0.720] to 0.737 [0.674-0.797]). Performance improved for most antibiotics when species data were included as model inputs (AUCs 0.723 [0.652-0.791] to 0.827 [0.797-0.857]). In patients treated with a beta-lactam, clinician prescribing led to 70% receiving an active beta-lactam: 44% were over-treated (broader spectrum treatment than needed), 26% optimally treated (narrowest spectrum active agent), and 30% under-treated (inactive beta-lactam). Model predictions without species data could have led to 79% of patients receiving an active beta-lactam: 45% over-treated, 34% optimally treated, and 21% under-treated.

**Interpretation:** Predicting AMR in bloodstream infections is challenging for both clinicians and models. Despite modest performance, machine learning models could still increase the proportion of patients receiving active empirical treatment by up to 9% over current clinical practice in an environment prioritising antimicrobial stewardship.

**Funding:** National Institute of Health Research (NIHR) Oxford Biomedical Research Centre, NIHR Health Protection Research Unit in Healthcare-associated Infection and Antimicrobial Resistance.

**Research in context:** 

**Evidence before this study:** We searched Pubmed and Google Scholar using the terms: [antibiotic OR antimicrobial] AND [resistance] AND [prediction OR machine learning OR AI OR artificial intelligence] for articles published up to 31 August 2024. References and citations for articles identified were also reviewed. Several studies have shown that machine learning can potentially be used to predict antimicrobial resistance (AMR) subsequently identified on phenotypic antimicrobial susceptibility testing. Most have focused either on identifying resistance in urinary tract infection, or in all samples received by a microbiology laboratory, which are often dominated by urine cultures. Only two studies were identified focusing specifically on bloodstream infection, and these only investigated a limited number of antibiotics. Overall, prediction performance was typically modest, e.g. area under the receiver operating curve (AUC) values of 0.65-0.75. Most studies focus on data available in the community or hospital but not both. Four studies retrospectively compared clinical prescribing to model predictions and showed models could potentially reduce inappropriate antibiotic use, but none focused specifically on bloodstream infection. External validation of models is uncommon, and most studies do not cover how models can be updated over time or to new locations.

**Added value of this study:** We developed machine learning models to predict resistance to seven antibiotics (amoxicillin, co-amoxiclav, ceftriaxone, piperacillin-tazobactam, ciprofloxacin, co-trimoxazole, and gentamicin) in bloodstream infections caused by Enterobacterales species. We focused on this clinical syndrome as it is an important cause of AMR-associated mortality. We used data from Oxfordshire, UK, between January 2017 and December 2023 for model training and evaluation (4709 infection episodes in 4243 patients). In held-out test data, predictive performance was similar for the seven antibiotics (AUCs 0.680 [95%CI 0.641-0.720] to 0.737 [0.674-0.797]). Performance improved for most antibiotics when species data were included as model inputs (AUCs 0.723 [0.652-0.791] to 0.827 [0.797-0.857]). AMR identified in recent microbiology results was the most important predictor of resistance. Model performance was relatively consistent over time. AMR prediction was also challenging for clinicians: their implied sensitivity for detecting resistance, i.e., the proportion of patients treated with a beta-lactam with resistance receiving active treatment was 97% for amoxicillin, 29% for co-amoxiclav, 19% for ceftriaxone, and 6% for piperacillin-tazobactam. In patients treated with a beta-lactam, clinician prescribing led to 70% receiving an active beta-lactam: 44% were over-treated (broader spectrum treatment than needed), 26% optimally treated (narrowest spectrum active agent), and 30% under-treated (inactive beta-lactam). Model predictions without species information could have led to 79% of patients receiving an active beta-lactam: 45% over-treated, 34% optimally treated, and 21% under-treated.

**Implications of all the available evidence:** Despite considering a wide range of input features, including hospital and some community data, model performance was broadly consistent with what has been described previously for similar tasks. This suggests there is a potential ceiling on the performance of machine learning in this context. However, despite modest performance, machine learning models could still increase the proportion of patients receiving active treatment by up to 9% over current clinical practice in an environment prioritising antimicrobial stewardship.

## Introduction

Active and timely antibiotic treatment of severe bacterial infections potentially saves lives and improves patient outcomes.[1] However, it can take up to 24-48 hours or more to obtain microbiology results to guide treatment, and many important infections may remain culture negative.[2] Therefore, substantial reliance is placed on antibiotic guidelines that are designed to maximise active empirical treatment of infections before microbiology results are available, while also minimising overuse of broad-spectrum antibiotics to avoid driving antimicrobial resistance (AMR).

Population-level antibiotic recommendations can be refined for individual patients, e.g. considering previous resistance or prior antibiotic exposure. However, this does not happen consistently, e.g. due to limited time available to retrieve earlier results or variable prescriber experience. Therefore, several previous studies have evaluated whether combining electronic healthcare record (EHR) data with predictive algorithms could improve detection of AMR and hence lead to better targeted prescribing (Table 1).[3–18] These studies typically focus on patients with positive microbiology and use machine learning to predict resistance to key antibiotics. Most previous studies focus on urinary tract infections or all infections (likely dominated by urine cultures), in part due to availability of large datasets for model training.[5–18] Only a minority focus specifically on bloodstream infection despite its clinical importance.[3,4] Several data types have been shown to be potentially informative, including a history of isolates with AMR, population AMR rates, previous personal antimicrobial exposure, past medical history and demographics. Data are typically obtained from a single hospital or community setting, but occasionally from a whole healthcare network.

**Table 1.** Previous models for predicting antibiotic resistance. Results shown are from an illustrative literature review. Only four studies were identified that have made their code publicly available.[10,15,17,18]

| Publication | Population | Infection type | Prediction input contains species information | Antibiotics | Model architecture | Features | Performance-AUC | External validation | Comparison with clinicians | Ref. |
| --- | --- | --- | --- | --- | --- | --- | --- | --- | --- | --- |
| Vazquez-Guillamet et al. 2017 | US, 2008-2015, single Hospital, 1618 patients | Bloodstream infection | Yes | Piperacillin-tazobactam (PT), cefepime (CE), meropenem (ME) | Logistic regression (LR), decision trees (DT) | Nursing home residence, transfer from an outside hospital, prior antibiotic use, source of infection, bacterial species | LR: 0.68, 0.63, and 0.83 for resistance to PT, CE, and ME<br>DT: 0.67, 0.61, and 0.80 for resistance to PT, CE, and ME | No | No | [3] |
| Sousa et al. 2019 | Spain, 2015-2016, single hospital, 448 samples | Gram negative bacteraemia | No | β-lactamase production | Decision tree | Comorbidities, source of infection, history of infection, antibiotic exposure, previous hospitalisation | 0.76 | No | No | [4] |
| Yelin et al. 2019 | Israel, 2007-2017, community health maintenance organization, 315,047 patients | Urinary tract infection | No | Co-trimoxazole, ciprofloxacin, co-amoxiclav, cefuroxime, cephalexin, nitrofurantoin | Logistic regression, gradient-boosted decision trees | Resistance profile, demographics, sample history, drug purchase history, cross-resistance | 0.70 (co-amoxiclav) to 0.83 (ciprofloxacin) | No | Yes | [5] |
| Fretzakis et al. 2020 | Greece, 2017-2018, single hospital, ICU patients, 345 patients | All infections | Partial (Gram stain) | Multiple | Random forest (RF), multi-layer perceptron (MLP) | Demographics, type of sample, Gram stain, antibiotics, previous antibiotic susceptibility testing | RF: 0.70<br>MLP: 0.73 | No | No | [6] |
| Hebert et al. 2020 | US, 2011-2016, single hospital, ICU patients, 6366 patients | Urinary tract infection | No | Cefazolin, ceftriaxone, ciprofloxacin, cefepime, and piperacillin-tazobactam | Logistic regression | Demographics, comorbidity score, recent antibiotic use, recent antimicrobial resistance, and antibiotic allergies | 0.65 (ceftriaxone) to 0.69 (cefazolin) | No | No | [7] |
| Lewin-Epstein et al. 2020 | Israel, 2013-2015, single hospital, 16,198 samples | All infections | No and Yes | Ceftazidime, gentamicin, imipenem, ofloxacin, | Logistic regression, neural networks, | Bacteria species, previous resistance, demographics, comorbidities, prior | Without species: 0.73–0.79<br>With species: 0.80–0.88 | No | No | [8] |
|  |  |  |  | sulfamethoxazole-trimethoprim | gradient boosted decision trees | hospitalisation, department/ward, previous antibiotics exposure |  |  |  |  |
| Moran et al. 2020 | UK, 2010-2016, 3 hospitals, 15,695 admissions | Community-associated bloodstream and urinary tract infections | No | Co-amoxiclav, piperacillin-tazobactam | XGBoost | Comorbidities, demographics, previous resistance, previous antibiotics exposure | 0.70 | No | Yes | [9] |
| Kanjilal et al. 2020 | US, 2007-2016, 2 hospitals, 13,682 patients, female, 18-55 years | Urinary tract infection | No | Nitrofurantoin, co-trimoxazole, ciprofloxacin, levofloxacin | Logistic regression, decision tree, random forest | Demographics, comorbidities, previous hospitalisation, previous procedures, lab tests, previous antibiotics use, previous resistance | Full cohort: 0.56-0.64; limit to prior antibiotic resistance or exposure: 0.61-0.77 | No | Yes | [10] |
| McGuire et al. 2021 | USA, 2012-2017, single hospital, 68,472 samples | All infections | No | Carbapenem | XGBoost | Demographics, medications, vital signs, prior procedures, lab tests, billing code, culture, sensitivity | 0.846 | No | No | [11] |
| Pascual-Sánchez et al. 2021 | Madrid, Spain, 2004-2020, single hospital, 3500 patients | All infections | No | Multiple, predict multi-drug resistance | Logistic regression, XGBoost, neural network, random forest | Time to culture, previous resistance | 0.76 | No | No | [12] |
| Martínez-Agüero et al. 2022 | Madrid, Spain, 2004-2020, single hospital, 3470 patients | All infections | No | Multiple agents | Long short-term memory network | Clinical time-series data | 0.67 | No | No | [13] |
| Rich et al. 2022 | US, 2011-2019, multi-centre, 6307 patients | Urinary tract infection | No | Co-trimoxazole (SXT), nitrofurantoin (NIT), ciprofloxacin (CIP), multi-drug resistance | Boosted logistic regression | Demographics, zip code, comorbidities, previous resistance, previous antibiotics exposure, previous hospital stay | 0.58 (SXT), 0.62 (NIT), 0.64 (CIP), and 0.66 (MDR) | Yes | No | [14] |
| Corbin et al. 2023 | US, 2009-2021, multi-centre, 8342 infections from 6920 patients | All infections | No | Vancomycin, piperacillin/tazobactam, cefepime, ceftriaxone, cefazolin, ciprofloxacin, ampicillin and meropenem | Gradient boosted decision tree, random forest | Diagnostic codes, prior procedures, lab tests, medications, respiratory care, previous resistance, vital signs, demographics, insurance, imaging, institution | 0.61-0.73 | Yes | Yes | [15] |
| Lee et al. 2023 | Korea, 2020-2021, single hospital, 550 samples | Urinary tract infection | No | Ciprofloxacin (CIP), extended-spectrum beta-lactamases (ESBL) | Gradient-boosted decision trees | Demographics, medical device, infection type, comorbidities, past history, vital signs, lab tests | 0.827 for CIP<br>0.811 for ESBL | No | No | [16] |
| Mintz et al. 2023 | Israel, 2016-2019, single hospital, 10053 samples | All infections | No and Yes | Ciprofloxacin | Super learner | Demographics, comorbidities, previous resistance, previous antibiotics exposure, department/ward | Without species: 0.737<br>With species: 0.837 | No | No | [17] |
| Yang et al. 2023 | US, 2007-2016, two hospitals, 101,096 samples | Urinary tract infection (UTI) | No | Nitrofurantoin (NIT), co-trimoxazole (SXT), ciprofloxacin (CIP), levofloxacin (LVX) | TabNet, XGBoost | Department/ward, demographics, previous resistance, previous organism, previous antibiotics exposure, comorbidities, previous procedures, colonisation pressure | Complicated UTI: 0.686 (NIT), 0.701 (SXT), 0.811 (CIP), 0.814 (LVX)<br>Uncomplicated UTI: 0.559 (NIT), 0.591 (SXT), 0.646 (CIP), 0.639 (LVX). | Validated on uncomplicated UTI | No | [18] |

In previous studies predictive performance for detecting AMR has been relatively modest, e.g. area under the receiver operating curve (AUC) values for important pathogen-antibiotic combinations of around 0.65-0.75, but varying between drugs and settings. If species identification is included as a model input performance improves, e.g. AUCs of 0.80-0.88.[8,17] However species is unknown when starting empirical treatment. Most studies use test data from the same setting either randomly chosen from the same period or from shortly after the training period, limiting generalisability over geographic locations and time. Within the 16 previous studies identified, only two externally validated their findings using data from a different area/hospital.[14,15] Most approaches do not address how to update models over time. Four studies retrospectively compared their model performance to clinical decision making, showing models could potentially reduce inappropriate antibiotic treatments.[5,9,10,15] Taken together, alongside technical barriers to interfacing with EHR systems and implementing models in healthcare settings, to date uptake of such predictions into clinical practice has been very limited.

Here we apply machine learning predictions to an important, but only partially studied patient group at particular risk poor outcomes from AMR, those with Enterobacterales bloodstream infection.[19] Our models are designed to be used in patients with suspected bloodstream infection where Enterobacterales are the most probable cause, e.g. infections with urinary or intra-abdominal focus. We use a comprehensive input feature set addressing potential limitations of some earlier studies, by combining data from hospital EHRs with community microbiology results. We also evaluate how performance changes over time and test approaches for updating models over time as new data emerges. We describe how good clinicians are at detecting AMR and compare the performance of our models to actual prescribing and simulate the impact that a prediction system might have on the number of patients receiving active antibiotic treatment, and the wider impact on use of broad-spectrum antibiotics.

## Methods

### Study design and population

We used data from Oxford University Hospitals (OUH), four teaching hospitals collectively providing 1100 beds, serving 750,000 residents in Oxfordshire, ∼1% of the UK population. The hospital’s microbiology laboratory also provides nearly all community testing for the region. Deidentified data were obtained from Infections in Oxfordshire Research Database (IORD), which has approvals from the South Central-Oxford C Research Ethics Committee (19/SC/0403), the Health Research Authority and the Confidentiality Advisory Group (19/CAG/0144) as a deidentified database without individual consent.

We included all patients aged ≥16 years with a positive blood culture containing a single Enterobacterales species between 01-January-2017 and 31-December-2023. Polymicrobial blood cultures were excluded as these potentially contained non-Enterobacterales species. Patients were included once per positive blood culture episode, i.e. including the first positive blood culture with an Enterobacterales species per 14-day period.

### Antimicrobial resistance prediction

We predicted antimicrobial susceptibility results for intravenous treatments for bloodstream infection that were commonly used in our institution with resistance rates >5%, i.e. amoxicillin, co-amoxiclav (amoxicillin-clavulanate), ceftriaxone, piperacillin-tazobactam, co-trimoxazole (trimethoprim-sulfamethoxazole), and ciprofloxacin. Predictions were not made for meropenem as resistance rates were <1%. Predicted results were binary, i.e. susceptible (including intermediate/dose-dependent susceptible) or resistant. During the study period, hospital empirical antibiotic guidelines recommended co-amoxiclav with or without additional single dose gentamicin for treatment of suspected sepsis of an unknown, urinary, or intra-abdominal source.

We made predictions at two time points, firstly at the time the blood culture was obtained and secondly when the species was identified. Input features included patient demographics, comorbidities, previous hospital-prescribed antibiotics, current clinical syndrome, previous specific AMR infections, the hour of day the blood culture was taken, counts of recent laboratory blood tests, previous hospital and community microbiology results including numbers of samples taken, number culture positive, and presence of antibiotic resistance to specific antibiotics, patient height and weight, previous hospital exposure, previous hospital-based procedures, current specialty, and counts of recent vital signs (Table S1). We also included recent population-level rates of AMR. The species-level analysis additionally included the species identified and any history of AMR in previous isolates of the same species. Overall, there were 152 features in the baseline model, and 182 in the species model.

### Model architecture, data partitioning and evaluation

We fitted separate XGBoost models for each antibiotic, aiming to predict subsequently identified phenotypic resistance. We used a temporal training-test split to mimic real-world implementation (training: 01-January-2017 to 31-December-2021; testing 01-January-2022 to 31-December-2022 (Test dataset 1)), reporting performance in the test dataset (details in supplement).

### Model updating

We used additional test data (Test dataset 2: 01-January-2023 to 31-December-2023) to evaluate if performance changed over time and different approaches for updating models. Three approaches were evaluated, in the first no further model training was undertaken, i.e. the model was based only on data from 2017-2021. In the second we retrained the model from scratch using all available data, i.e. from 2017-2022 inclusive. In the final approach we used on online-training method, where the trained model from the 2017-2021 was updated with data from 2022, using an inbuilt method available within XGBoost.

### Comparison with clinical decision making

To compare our models with clinical practice, we combined both test datasets and considered patients initially treated with a beta-lactam antibiotic. Beta-lactams were the most commonly used antibiotics in our institution and facilitated establishing a hierarchy of antibiotic choices. We included patients empirically treated with amoxicillin, co-amoxiclav, ceftriaxone, piperacillin-tazobactam, or a carbapenem (mostly meropenem; a small number receiving empirical ertapenem), in order of increasing spectrum of coverage. The most common adjunctive antibiotic in our setting was single dose gentamicin, however we exclude it here from our main analysis considering only the beta-lactam given, as we have previously shown gentamicin does not rescue patients with beta-lactam (co-amoxiclav) resistance from associated increases in mortality in *Escherichia coli* bloodstream infection.[20] We excluded from the clinical comparison neutropenic patients, patients not started on antibiotics, and blood cultures missing one or more susceptibility results for the beta-lactams listed above. No patient allergy data were available.

To compare clinical practice and models predictions, we evaluate the number of patients who are i) optimally treated, i.e. received the least broad-spectrum beta-lactam to which their blood culture isolate is sensitive, ii) under-treated, given a beta-lactam with resistance present, and iii) over-treated, given an active beta-lactam, but one that was of a broader spectrum than was necessary. We also describe the relative usage rates of each antibiotic.

We evaluated 4 strategies for applying our machine learning predictions without species information, tuning the prediction thresholds using the training data to: 1) match total antibiotic use to total clinician antibiotic use, but distributing it between patients more optimally, 2) match total use to population antibiotic susceptibility rates, 3) to match rates of over-treatment by clinicians, while aiming to increase active treatment, and 4) to see how much our models could reduce over-treatment if the default antibiotic policy was switched from using co-amoxiclav to ceftriaxone first-line (details in Supplement). Thresholds were then applied in the combined test data and performance summarised.

## Results

Between 01-January-2017 and 31-December-2023, 252,849 blood cultures were obtained. 24,228 (9.6%) were culture positive, including 6983 (2.8%) with an Enterobacterales species. After removing polymicrobial infections and de-duplicating repeat positive samples within 14 days, there were 4752 infection episodes in 4273 patients, a further 43 blood cultures were excluded because antimicrobial susceptibility testing was not performed, leaving 4709 infection episodes in 4243 patients for model training and evaluation (Figure 1). The median (IQR) patient age was 74 (60-84) years, and 2611 (55%) episodes were in male patients. The median (IQR) Charlson co-morbidity score was 1 (0-3). 3631 (77%) of positive blood cultures were community onset (i.e. within <48 hours of hospital admission).

**Figure 1.**
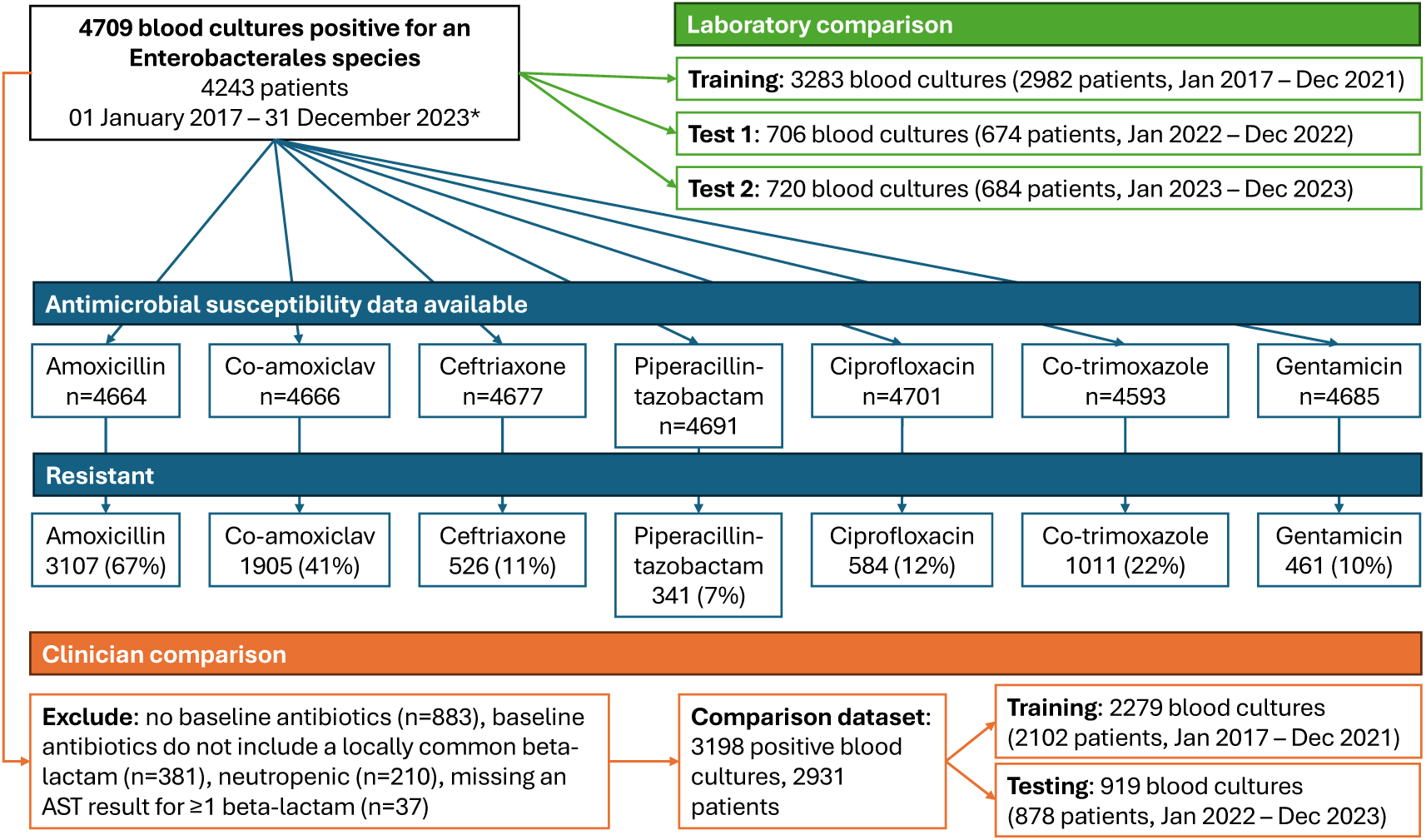
Blood cultures studied, and laboratory and clinical comparison groups. Repeat positive cultures from the same patient within the next 14 days after a positive blood culture were excluded. Only 7 blood cultures were resistant to meropenem, in the laboratory comparison 3 were in the training data, 2 in test 1 and 2 in test 1; in the clinical comparison 3 meropenem resistant blood cultures were included, 1 in the training data and 2 in the test data. Not all blood cultures had susceptibility results reported for all antibiotics as shown. Within the 3198 blood cultures studied in the clinical comparison, 2064 (65%) were resistant to amoxicillin, 1225 (38%) to co-amoxiclav, 320 (10%) to ceftriaxone, 190 (6%) to piperacillin-tazobactam, and 3 (<1%) to meropenem. Rates of resistance to gentamicin, ciprofloxacin and co-trimoxazole were 318/3195 (10%), 379/3196 (12%), and 674/3170 (21%) respectively.

The most commonly isolated species were *E. coli* (3094, 66%), *Klebsiella pneumoniae* (545, 12%), *Proteus mirabilis* (203, 4%), *Enterobacter cloacae* (177, 4%), and *K. oxytoca* (153, 3%). Across all species 3107/4664 (67%) were resistant to amoxicillin, 1905/4666 (41%) to co-amoxiclav, 526/4677 (11%) to ceftriaxone, 341/4691 (7%) to piperacillin-tazobactam, 461/4685 (10%) to gentamicin, 1011/4593 (22%) to co-trimoxazole, and 584/4701 (12%) to ciprofloxacin (denominator varies as not all samples were tested for all antibiotics). Resistance to meropenem was uncommon, 7/4689 (<0.2%).

The most frequently prescribed empirical antibiotics given within 4 hours of obtaining the blood cultures were co-amoxiclav alone (1194, 25%), no antibiotics (883, 19%), co-amoxiclav + gentamicin (761, 16%), ceftriaxone alone (236, 5%), and piperacillin-tazobactam alone (109, 2%); 82 (2%) patients received a carbapenem with or without another antimicrobial.

### Model performance at baseline

In held-out test data from 2022 (Test dataset 1), predictive performance was broadly similar for the seven antibiotics, AUCs ranging from 0.680 [95%CI 0.641-0.720] for amoxicillin to 0.737 [0.674 - 0.797] for ceftriaxone (Table 2; Table S4 for training data performance). Jointly optimising sensitivity and specificity, sensitivity ranged from 40.7% (32.7-49.6%) for co-trimoxazole to 62.2% (57.8-66.6%) for amoxicillin, while specificity ranged from 66.4% (60.2-72.3%) for amoxicillin to 91.5% (89.2-93.7%) for co-trimoxazole. Positive predictive values (PPVs) and negative predictive values (NPVs), which are influenced by differences in resistance prevalence, ranged from 19.8% (14.2-26.3%) to 78.0% (73.9-81.9%) and 47.9% (42.6-53.7) to 94.2% (92.2-96.0%), respectively. Alternative values for sensitivity/specificity/PPV/NPV could be obtained by varying the threshold chosen for identifying resistance (e.g. prioritising sensitivity, Table S5).

**Table 2.** Model performance for predicting antibiotic resistance at blood culture sampling in held-out test dataset 1, 01 January 2022 – 31 December 2022. AUC, area under the receiver operating curve. Confidence intervals were generated by bootstrapping with 1000 iterations.

| Antibiotic | n | Resistant,<br>n | Resistant,<br>% | AUC (95%CI) at blood<br>culture sampling | Sensitivity<br>(95% CI) | Specificity<br>(95% CI) | Positive predictive<br>value (95% CI) | Negative predictive<br>value (95% CI) |
| --- | --- | --- | --- | --- | --- | --- | --- | --- |
| Amoxicillin | 693 | 455 | 66 | 0.680 (0.641 - 0.720) | 62.2 (57.8 - 66.6) | 66.4 (60.2 - 72.3) | 78.0 (73.9 - 81.9) | 47.9 (42.6 - 53.7) |
| Co-amoxiclav | 699 | 283 | 40 | 0.684 (0.642 - 0.722) | 60.4 (54.7 - 66.1) | 67.8 (63.7 - 71.9) | 56.1 (50.5 - 61.3) | 71.6 (67.8 - 75.8) |
| Ceftriaxone | 701 | 78 | 11 | 0.737 (0.674 - 0.797) | 48.7 (37.5 - 60.2) | 83.0 (80.1 - 85.9) | 26.4 (19.2 - 34.2) | 92.8 (90.6 - 94.9) |
| Piperacillin-tazobactam | 704 | 64 | 9 | 0.708 (0.643 - 0.779) | 51.6 (39.6 - 63.8) | 79.1 (75.8 - 82.1) | 19.8 (14.2 - 26.3) | 94.2 (92.2 - 96.0) |
| Ciprofloxacin | 706 | 86 | 12 | 0.726 (0.655 - 0.789) | 50.0 (39.2 - 60.6) | 88.2 (85.7 - 90.6) | 37.1 (28.0 - 45.4) | 92.7 (90.6 - 94.8) |
| Co-trimoxazole | 688 | 123 | 18 | 0.698 (0.641 - 0.754) | 40.7 (32.7 - 49.6) | 91.5 (89.2 - 93.7) | 51.0 (41.7 - 60.5) | 87.6 (84.8 - 90.3) |
| Gentamicin | 704 | 75 | 11 | 0.700 (0.625 - 0.775) | 45.3 (34.2 - 57.1) | 85.2 (82.6 - 88.0) | 26.8 (19.4 - 35.2) | 92.9 (90.6 - 94.8) |

### Model performance following species identification

Performance improved for most antibiotics when species data were included as inputs to the prediction models, i.e., mimicking the point during laboratory work-up of a blood culture when the species is first identified, but susceptibility results remain pending. For example, AUCs increased for amoxicillin (0.680 [95%CI 0.641-0.720] to 0.827 [0.797-0.857]) and co-amoxiclav (0.684 [0.642-0.722] to 0.771 [0.734-0.805]). Performance increases were also seen for other antibiotics, but with minimal improvement for piperacillin-tazobactam (Table 3, Table S6 for training dataset).

**Table 3.** Model performance for predicting antibiotic resistance at blood culture species identification in held-out test dataset 1, 01 January 2022 – 31 December 2022. AUC, area under the receiver operating curve. Confidence intervals were generated by bootstrapping with 1000 iterations.

| Antibiotic | n | Resistant,<br>n | Resistant,<br>% | AUC (95%CI) with<br>species information | Sensitivity<br>(95% CI) | Specificity<br>(95% CI) | Positive predictive<br>value (95% CI) | Negative predictive<br>value (95% CI) |
| --- | --- | --- | --- | --- | --- | --- | --- | --- |
| Amoxicillin | 693 | 455 | 66 | 0.827 (0.797 - 0.857) | 65.1 (60.8 - 69.9) | 85.7 (81.3 - 90.0) | 89.7 (86.4 - 92.8) | 56.2 (51.3 - 61.5) |
| Co-amoxiclav | 699 | 283 | 40 | 0.771 (0.734 - 0.805) | 67.1 (61.0 - 72.6) | 72.8 (68.3 - 77.1) | 62.7 (57.3 - 68.1) | 76.5 (72.4 - 80.7) |
| Ceftriaxone | 701 | 78 | 11 | 0.799 (0.745 - 0.846) | 64.1 (53.1 - 73.9) | 77.5 (74.1 - 80.9) | 26.3 (19.8 - 32.4) | 94.5 (92.5 - 96.3) |
| Piperacillin-tazobactam | 704 | 64 | 9 | 0.723 (0.652 - 0.791) | 64.1 (51.9 - 75.4) | 67.3 (63.6 - 71.1) | 16.4 (12.1 - 20.8) | 94.9 (92.7 - 96.8) |
| Ciprofloxacin | 706 | 86 | 12 | 0.783 (0.724 - 0.840) | 47.7 (37.1 - 59.2) | 92.7 (90.6 - 94.7) | 47.7 (36.6 - 57.8) | 92.7 (90.7 - 94.8) |
| Co-trimoxazole | 688 | 123 | 18 | 0.774 (0.726 - 0.821) | 49.6 (41.3 - 58.9) | 89.7 (87.2 - 92.2) | 51.3 (42.6 - 60.2) | 89.1 (86.6 - 91.6) |
| Gentamicin | 704 | 75 | 11 | 0.729 (0.654 - 0.794) | 41.3 (29.5 - 52.6) | 91.7 (89.5 - 93.9) | 37.3 (26.8 - 47.4) | 92.9 (90.9 - 94.8) |

### Feature importance

The most important features for making predictions were relatively consistent across different antibiotics (Figure 2, Figures S1-S6). The time since the last isolate with resistance to the specific antibiotic modelled was the most important feature for all antibiotics except piperacillin-tazobactam. Shorter times contributed most strongly to a prediction of resistance, with the importance of a previous resistant isolate to the same antibiotic typically attenuating over 1 year (Figure 3). Other consistently important features included greater time since hospital admission at blood culture sampling, shorter time since a previous resistant isolate to other related antibiotics, increased hospital antibiotic exposure (specifically for the antibiotic of interest, related antibiotics, and total antibiotics), and recent blood and urine cultures being sent.

**Figure 2.**
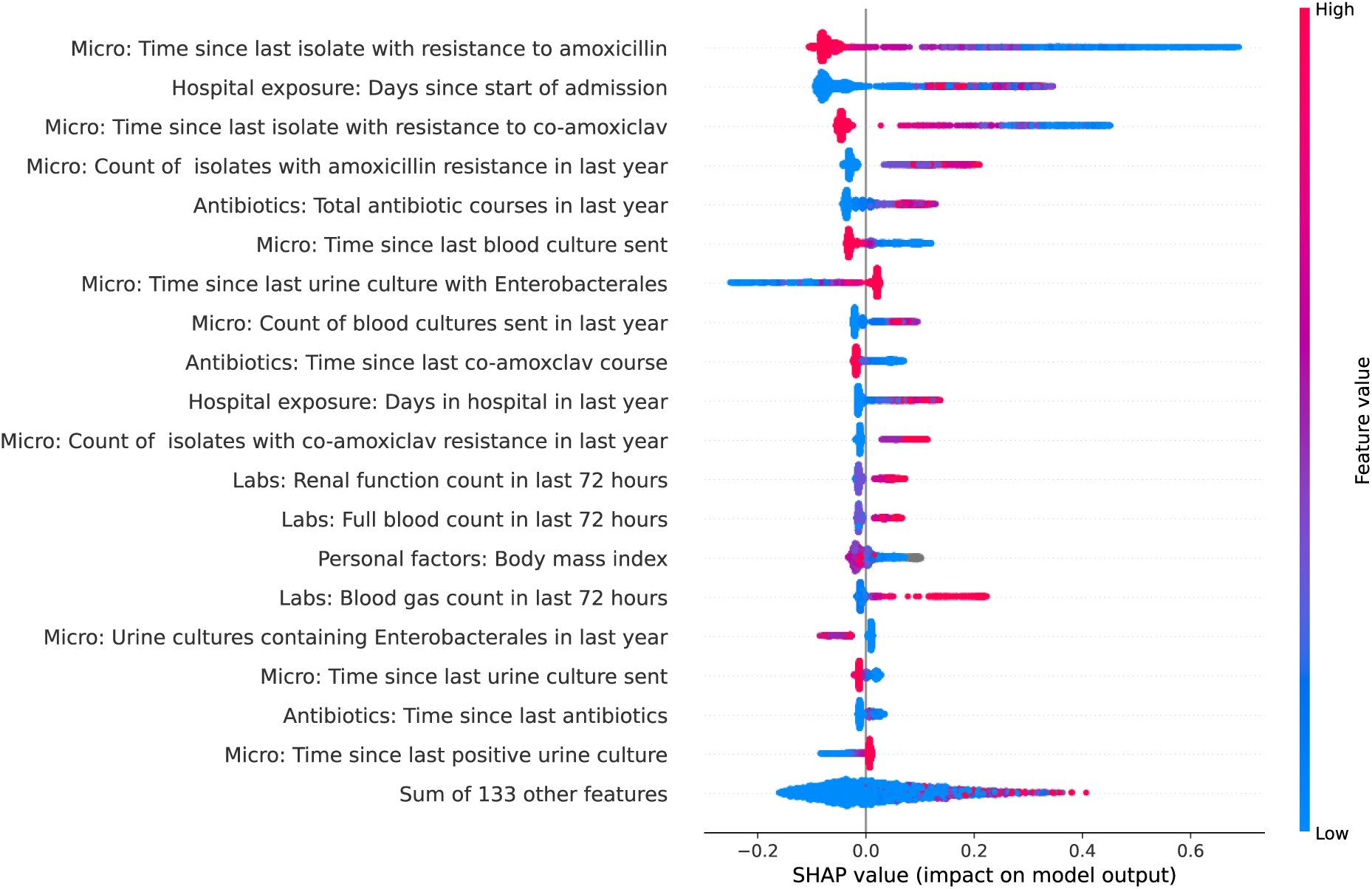
SHAP (SHapley Additive exPlanations) plot showing feature importance and impacts on model output for predicting amoxicillin resistance at blood culture sampling. Positive values on the x-axis indicate contributions towards predicting resistance, and negative values contributions towards predicting susceptibility. Absolute x-axis values reflect the relative importance or contribution of the feature in making a prediction. Colour indicates the value of the feature, red dots indicate higher values and blue dot lower values. For example, the shorter the time since the last isolate with resistance to amoxicillin the more likely a prediction of resistance. See Figures S1-S6 for other antibiotics. Shorter times since the last urine culture with Enterobacterales were associated with predicting susceptibility, although this might seem surprising, it needs to be interpreted considering also having the time since a resistant isolate in the model, such that given that result, shorter times may represent evidence of a recent susceptible isolate.

**Figure 3.**
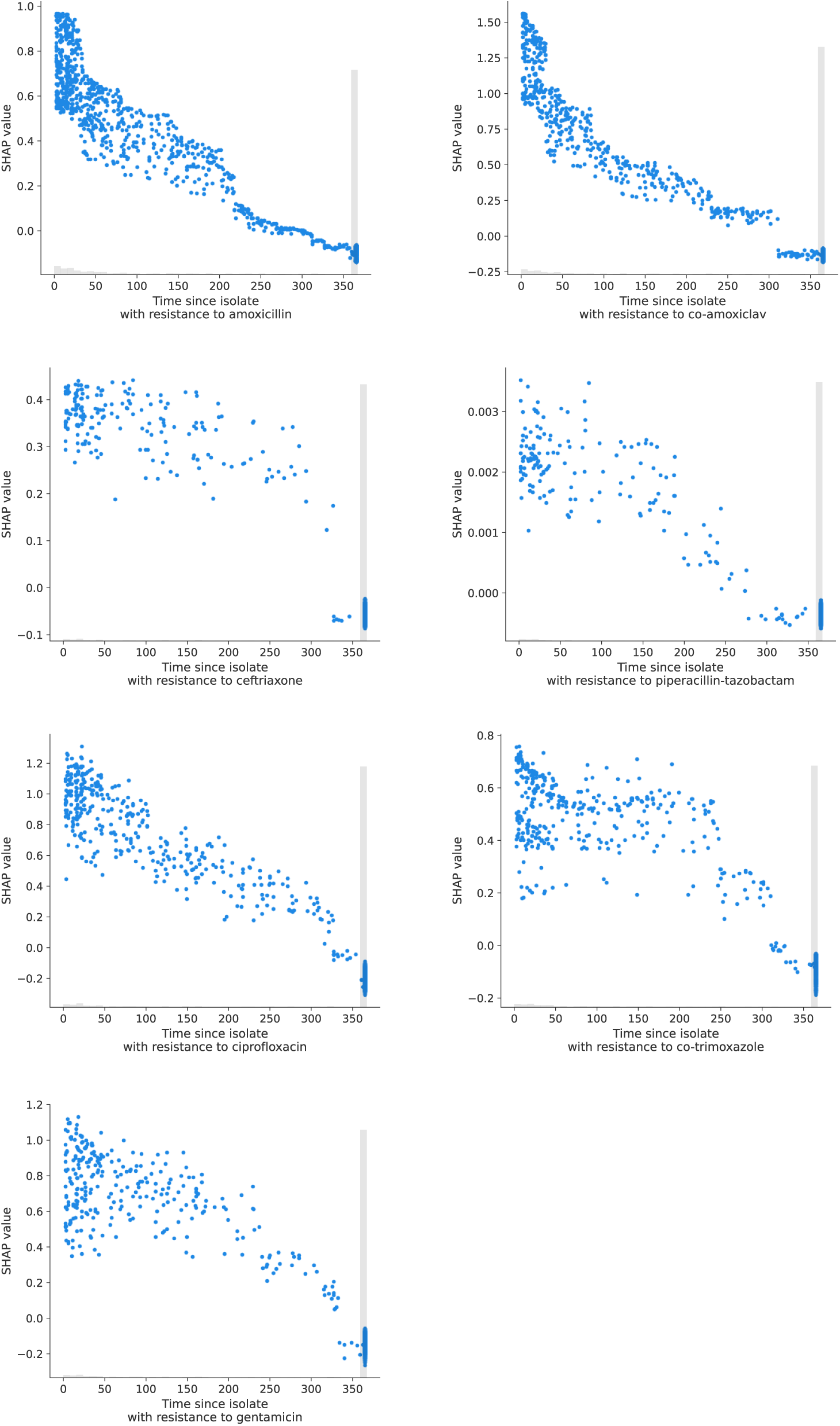
SHAP (SHapley Additive exPlanations) plots showing the time since last resistant isolate and impact on model output for predicting resistance to the same antibiotic at blood culture sampling. Where no resistant isolate was seen in the last year, the value is set to 365, hence when interpreting change over time values exactly equal to 365 days should be ignored. The grey histogram indicates the relative frequency of each observation on the x-axis. The spread of blue points arises from other features also influencing the SHAP value on the y-axis.

When the species identified was added as a model input, this also became an important model feature, particularly for antibiotics where species information improved model performance the most, such as amoxicillin and co-amoxiclav (Figure S7-13). In some cases, the features reflected information in the data arising from intrinsic resistance (e.g. *Klebsiella spp*. and amoxicillin), but in other cases reflected different resistance prevenances in different species.

### Model updates over time

Using held-out test data from 2023 (Test Dataset 2) there was minimal evidence that model performance changed over time compared to 2022 and with similar results in 2023 using the original model, a retrained model, and an incrementally updated model (Figure 4). Models were relatively quick to train, taking around a minute on a high-performance personal computer, such that savings in training time from not updating the models or from incremental updating were minimal (Figure S14).

**Figure 4.**
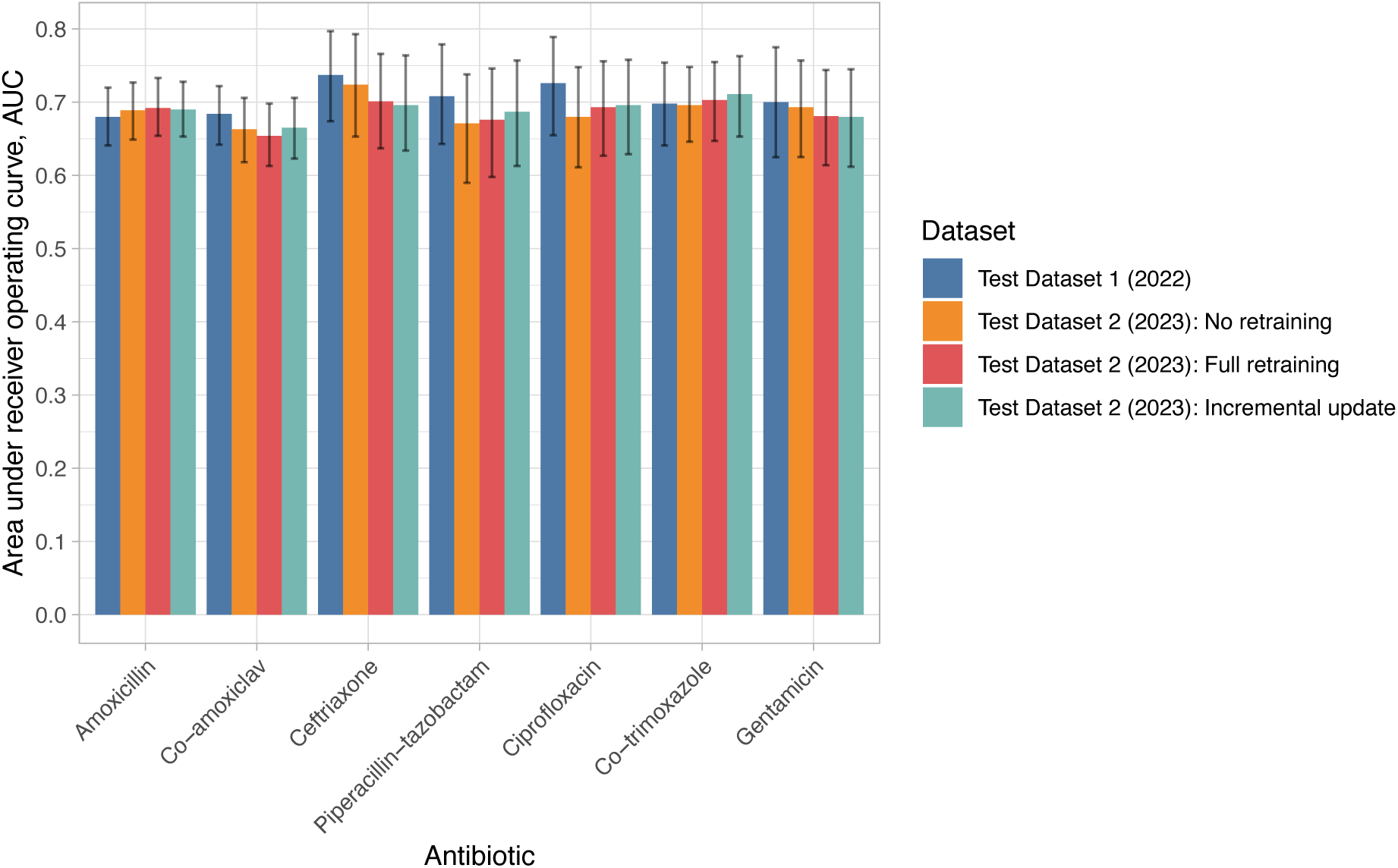
Model performance for predicting antibiotic resistance at blood culture sampling in held-out test dataset 1 (01 January 2022 – 31 December 2022) and 2 (01 January 2023 – 31 December 2023). For test dataset 2 three approaches to updating the model over time are presented – no retraining, full re-training from scratch using data from 2017-2022 inclusive, incremental updating of the original model trained using 2017-2021 data with the data from 2022. AUC, area under the receiver operating curve. Confidence intervals were generated by bootstrapping with 1000 iterations.

### Comparison to clinical practice

Of the 4709 blood cultures positive for an Enterobacterales species, 3198 were included in the clinical comparison analysis (see Figure 1 for exclusions).

#### Antibiotic use by clinicians – over, under and optimal treatment

Of the 3198 infections, 2512 (79%) received at least one active baseline antibiotic, and 686 (21%) did not. Considering the beta-lactam given, 806 (25%) were optimally treated, 974 (30%) under-treated, and 1418 (44%) were over-treated. Most patients were treated with co-amoxiclav (2286, 71%), which also accounted for the greatest proportion of patients under-treated (Figure 5A). In an ideal scenario where all infections were treated with the narrowest spectrum active antibiotic, more patients would have received amoxicillin, fewer co-amoxiclav and more ceftriaxone, with small increases compared to actual practice in piperacillin-tazobactam and carbapenem use too (Figure 5B). Most patients given inactive treatment would have been optimally treated with ceftriaxone with a smaller number requiring piperacillin-tazobactam or a carbapenem (Figure 5C).

**Figure 5.**
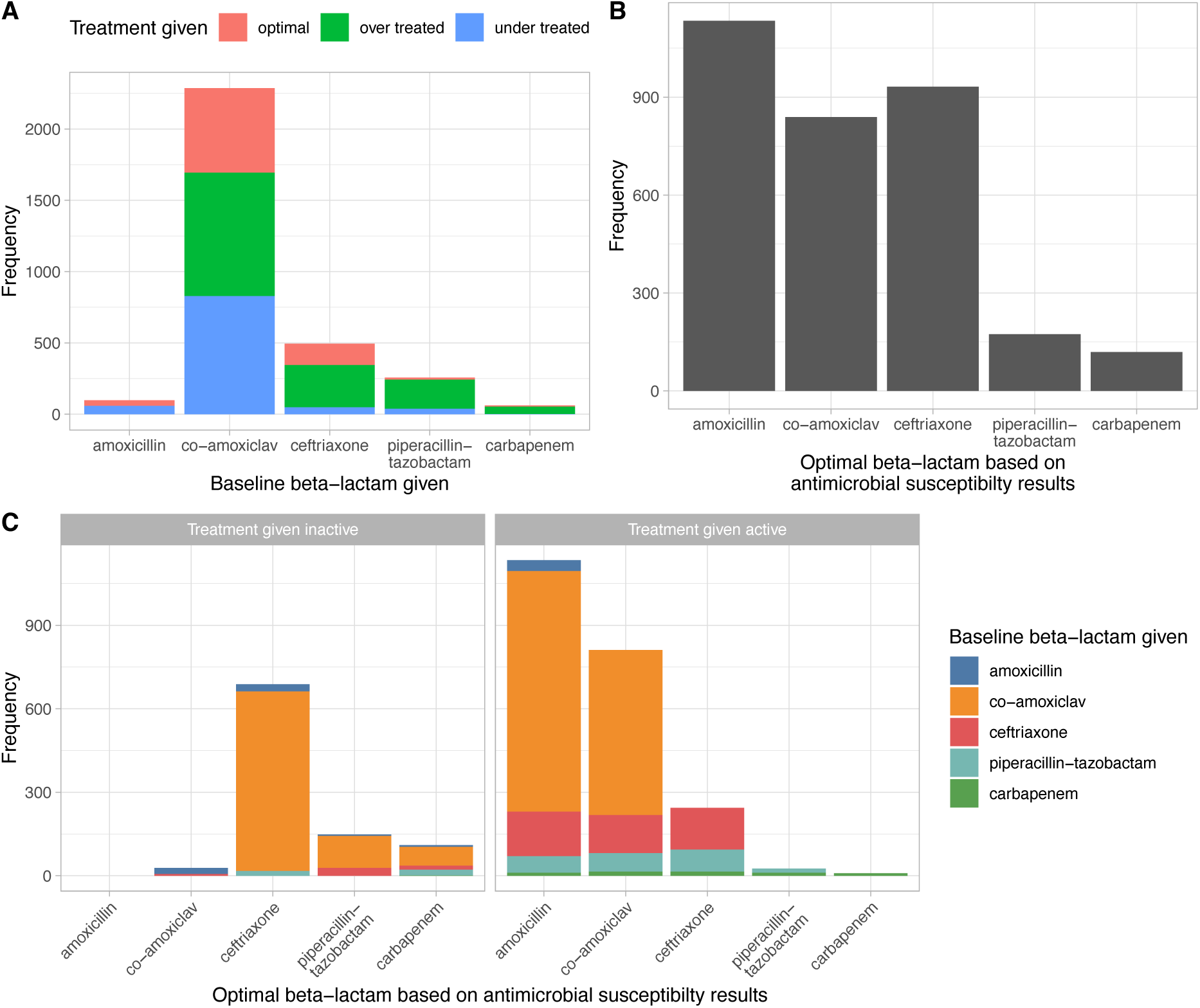
Clinician prescribing practice in 3198 positive blood cultures. Panel A shows the number of infections treated with different beta-lactams, classified by whether treatment was optimal, broader than necessary (‘over treated’), or had resistance to the beta-lactam used (‘under treated’). Panel B displays the optimal breakdown of antibiotic use, had the narrowest spectrum active agent been used to treat each infection. Panel C shows the distribution of optimal antibiotics by whether the actual beta-lactam treatment given was inactive (left hand sub-panel) or active (right).

The implied sensitivity of clinicians for detecting resistance, i.e., the proportion of patients with resistance to a given antibiotic receiving treatment with any active broader spectrum beta-lactam was 97% (2005/2064) for amoxicillin, 29% (360/1225) for co-amoxiclav, 19% (61/320) for ceftriaxone, and 6% (11/190) for piperacillin-tazobactam.

#### Strategy 1 – matching antibiotic use

Firstly, model performance without species information was compared to clinicians by attempting to constrain the predictions made to result in the same total number of prescriptions for each antibiotic as used by clinicians. Within the combined test data, clinician prescribing resulted in 70% of patients receiving an active beta-lactam: 44% were over-treated, 26% optimally treated, and 30% under-treated. Model predictions resulted in more patients being actively treated, 75%, fewer being over-treated, 42%, fewer under-treated, 25%, and therefore more being optimally treated, 33% (Figure 6A). Due to differences in model fit and calibration between training and test data small changes in antibiotic use were observed (Figure 6B, Table 4).

**Figure 6.**
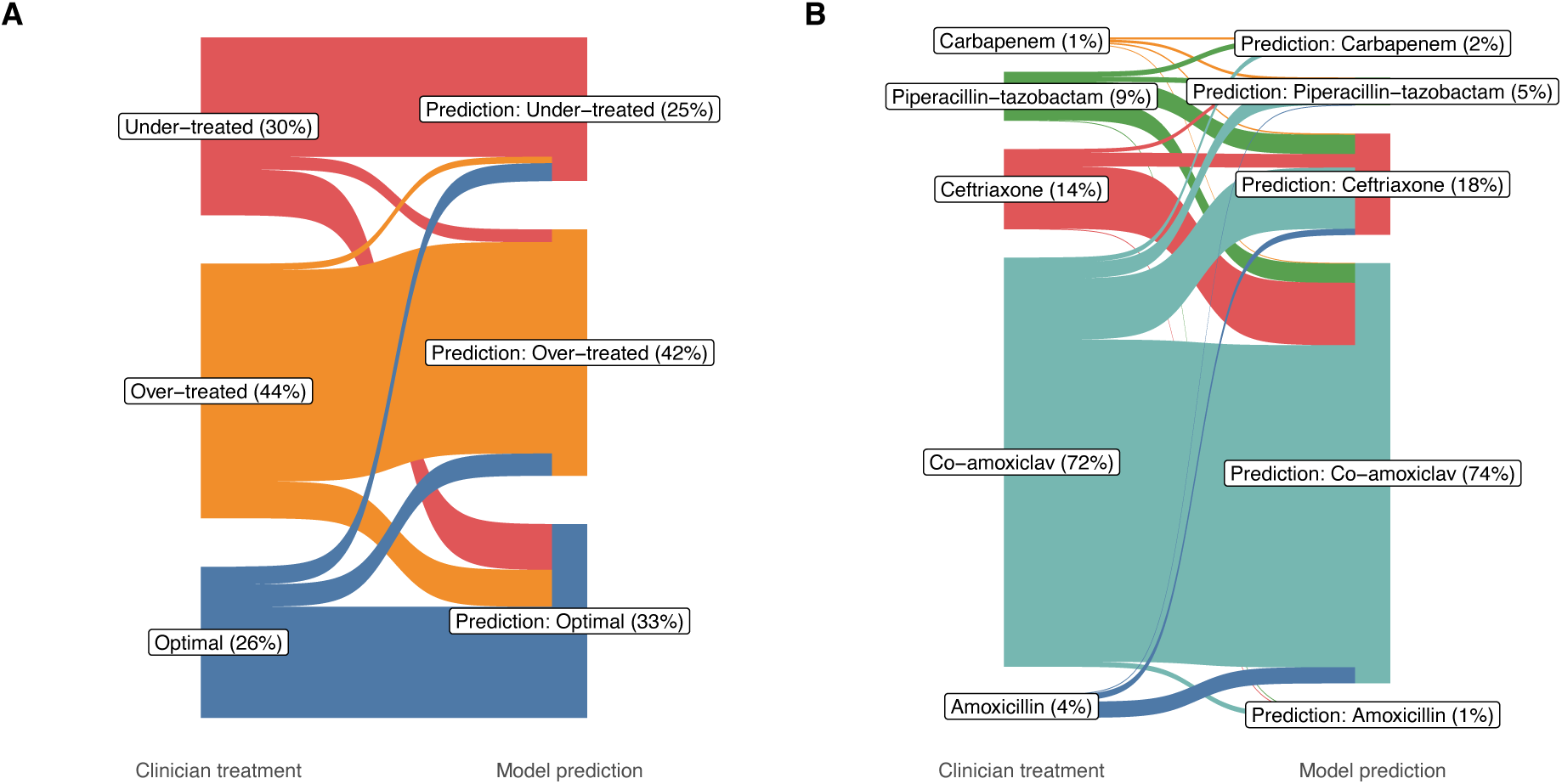
Percentage of patients receiving optimal, under or over treatment (panel A), and specific antibiotics (panel B) according to clinician prescribing and model predictions. Predictions in test data from 2022-2023 are shown for a model constrained to match the total use of each antibiotic as closely as possible (scenario 1) (differences in antibiotic use arise from differences between training and test dataset model fit and calibration).

**Table 4.** Comparison of model predictions to clinician prescribing in test data from 2022-2023 (n=919). Patients treated with a beta-lactam were included. Model scenarios are described in more details in the Methods.

| Scenario | Active beta-lactam, n (%) | Under-treated, n (%) | Optimally treated, n (%) | Over-treated, n (%) | Receiving amoxicillin, n (%) | Receiving co-amoxiclav, n (%) | Receiving ceftriaxone, n (%) | Receiving piperacillin-tazobactam, n (%) | Receiving carbapenem, n (%) |
| --- | --- | --- | --- | --- | --- | --- | --- | --- | --- |
| Clinician prescribing | 639 (70%) | 280 (30%) | 238 (26%) | 401 (44%) | 37 (4%) | 663 (72%) | 130 (14%) | 79 (9%) | 10 (1%) |
| Model, strategy 1: matching clinician antibiotic use | 693 (75%) | 226 (25%) | 305 (33%) | 388 (42%) | 10 (1%) | 681 (74%) | 164 (18%) | 45 (5%) | 19 (2%) |
| Model, strategy 2: matching antibiotic susceptibility rates | 655 (71%) | 264 (29%) | 384 (42%) | 271 (29%) | 320 (35%) | 238 (26%) | 301 (33%) | 23 (3%) | 37 (4%) |
| Model, strategy 2: matching antibiotic susceptibility rates with 20% leeway | 705 (77%) | 214 (23%) | 374 (41%) | 331 (36%) | 258 (28%) | 194 (21%) | 395 (43%) | 26 (3%) | 46 (5%) |
| Model, strategy 3: matching clinician over-treatment rates | 724 (79%) | 195 (21%) | 309 (34%) | 415 (45%) | 0 (0%) | 543 (59%) | 356 (39%) | 16 (2%) | 4 (<1%) |
| Simple comparator: switch first-line antibiotic to ceftriaxone | 817 (89%) | 102 (11%) | 221 (24%) | 596 (65%) | 0 (0%) | 0 (0%) | 830 (90%) | 79 (9%) | 10 (1%) |
| Model, strategy 4: matching clinician active treatment rates in ceftriaxone first-line comparator | 822 (89%) | 97 (11%) | 282 (31%) | 540 (59%) | 5 (1%) | 59 (6%) | 836 (91%) | 15 (2%) | 4 (<1%) |

#### Strategy 2 – matching antibiotic use to susceptibility rates

Using alternative prediction thresholds that matched total antibiotic use to susceptibility rates, resulted in more patients receiving optimal treatment compared to clinician’s prescribing, 42% (cf. 26% by clinicians) and reduced over-treatment in 29% (cf. 44%). However, overall active treatment was similar, 71% (cf. 70%). In a sensitivity analysis where 20% reductions in amoxicillin and co-amoxiclav use were allowed relative to susceptibility rates, the percentage of patients predicted to receive active treatment increased to 77%, while overtreatment at 36% remained less than that seen with clinician’s prescribing.

#### Strategy 3 – fixing overtreatment rates, aiming for more active treatment

If prediction thresholds were set by matching clinician over-treatment rates, 79% of patients would receive an active beta-lactam, 34% optimally treated, 45% over-treated, and 21% under-treated. This performance was predominantly achieved by moving a subset of patients treated by clinicians with co-amoxiclav to receive ceftriaxone, with fewer given piperacillin-tazobactam or a carbapenem to offset this.

#### Switching to ceftriaxone as first-line treatment and strategy 4

We also compared performance to a simpler intervention, changing first-line treatment to ceftriaxone, i.e. switching all patients receiving amoxicillin or co-amoxiclav to ceftriaxone. This would reduce the number of under-treated patients to 11%, but also cause 65% to be over-treated. Setting our models to achieve the same level of active treatment allowed over-treatment to be reduced to 59%.

## Discussion

Machine learning models can predict resistance to commonly used antimicrobials in Enterobacterales bloodstream infection with moderate accuracy. Despite considering a wide range of input features, including hospital and some community data, model performance was broadly consistent with what has been described previously for similar tasks.[3–18] This suggests there is a ceiling on the performance of machine learning in this context that is unlikely to be improved on without further data, e.g. for our models, data on community prescribing. It may also reflect intrinsic stochasticity where bacteria with and without AMR exist within a patient’s microbiome, and cause disease with or without AMR with a degree of randomness.

Despite modest performance of machine learning models, detecting resistance is also a highly challenging task for clinicians. In our hospital group antimicrobial stewardship is given a high priority, seeking to minimise over-use of broad-spectrum antibiotics to protect local needs and meet national prescribing incentives.[21,22] However, this also results in a substantial proportion of patients receiving inactive initial treatment, 21% overall, with 30% receiving inactive initial beta-lactam treatment. The implied sensitivity of clinician detection of resistance was low at 29% for co-amoxiclav, 19% for ceftriaxone, and 6% for piperacillin-tazobactam. Prescriptions made up to 4 hours after blood culture sampling were considered, such that they are likely to represent the practice of clinicians with a range of experiences, e.g., from 1-2 years post-graduation to more than 10 years, as senior reviews will not all have been completed for all patients within the time window chosen. However, this is representative of those making antibiotic prescribing decisions.

The challenging nature of antibiotic selection meant clinician’s choice of beta-lactam resulted in 30% of patients being under-treated, 44% being over-treated and only 26% being optimally treated. Several modelling approaches were able to improve on this. If total antibiotic use was kept broadly similar, but redistributed, an additional 5% of patients received active treatment, 75% overall, and optimal treatment rose to 33%. Alternatively, if we matched antibiotic use to rates of antibiotic susceptibility and allowed for some over-prescribing of broader spectrum agents to offset imperfect model performance 79% of patients could be actively treated, 9% more than by clinicians, while still only overtreating 45% (similar to clinicians). A simpler approach of switching all first-line antibiotics to ceftriaxone, increased active treatment to 89%, but caused 65% to be overtreated, the latter could be reduced to 59% if a model rather than a guideline change was used.

Model performance was relatively consistent over time and models could be retrained rapidly if needed. The features contributing most to predictions, such as infections with AMR within the last year or antibiotic exposures are likely to be relatively stable over time. To improve overall predictive performance and facilitate antimicrobial stewardship, better models are particularly needed for the narrower spectrum agents including amoxicillin and co-amoxiclav. Data on community use of these antibiotics and other narrow spectrum agents may help. It is unlikely that other model architectures would have substantially improved performance given the range of approaches tried in other studies without much better performance.[8,12,13,17,18]

This study has several limitations. A fundamental limitation with all studies of this kind is that the data are trained and tested on patients known to have positive blood cultures, and in our models, we specifically focused on patients with Enterobacterales species in blood cultures. However, <10% of sampled patients will have positive blood cultures and these results are not known *a priori*. Hence, we have to assume the predictors of resistance are similar in those with and without positive cultures, and that rates of resistance are similar too. However, this may only be partially true, especially if AMR contributes to blood cultures being positive in patients with prior antibiotic exposures in the community. By focusing on Enterobacterales we also did not consider infections with other species or polymicrobial infections. We did not have data available on allergies, it is possible that some of the model improvements in reducing over-treatment from switching ceftriaxone to co-amoxiclav may not have been possible due to penicillin allergies. However, this is less important for gains in the number of patients receiving active treatment, where the switches were generally to ceftriaxone. We did not have data on community antibiotic exposures, which may have improved model performance. We did however have data on community microbiology samples as nearly all samples were sent to a single hospital laboratory. It is possible that clinician prescribing in Oxfordshire is unusual in the priority given to antibiotic stewardship, but national prescribing data suggest it is not atypical for the UK.[22] In our setting patients receiving inactive initial treatment are typically switched to active treatment within 24-72 hours of blood cultures being obtained.[20] Our model was validated on two independent internal validation datasets, but further external validation is required before it can be deployed, for example as a decision support aid for hospital clinicians.

In conclusion, predicting who will have AMR is challenging for clinicians and models alike. Despite relatively modest performance of machine learning models, these could still increase the proportion of patients receiving active treatment by up to 9% over current clinical practice in an environment prioritising antimicrobial stewardship.

## Author contributions

DWE and ASW conceived the study. DWE and KY analysed the data. KY, AL and JW performed the literature review. Oversight was provided by DWE, TZ and ASW. DWE wrote the first draft of the manuscript assisted by KY, AL and JW. All authors revised the manuscript.

## Data Availability

The datasets analysed during the current study are not publicly available as they contain personal data but are available from the Infections in Oxfordshire Research Database (https://oxfordbrc.nihr.ac.uk/ research-themes-overview/antimicrobial-resistance-and-modernising-microbiology/infections-in-oxfordshire-research-database-iord/), subject to an application and research proposal meeting the ethical and governance requirements of the Database. For further details on how to apply for access to the data and for a research proposal template please.

## Acknowledgements

This work uses data provided by patients and collected by the UK’s National Health Service as part of their care and support. We thank all the people of Oxfordshire who contribute to the Infections in Oxfordshire Research Database. Research Database Team: L Butcher, H Boseley, C Crichton, DW Crook, D Eyre, O Freeman, J Gearing (public respresentative), R Harrington, K Jeffery, M Landray, A Pal, TEA Peto, TP Quan, J Robinson (public respresentative), J Sellors, B Shine, AS Walker, D Waller. Patient and Public Panel: G Blower, C Mancey, P McLoughlin, B Nichols.

## Data sharing

The datasets analysed during the current study are not publicly available as they contain personal data but are available from the Infections in Oxfordshire Research Database (https://oxfordbrc.nihr.ac.uk/research-themes-overview/antimicrobial-resistance-and-modernising-microbiology/infections-in-oxfordshire-research-database-iord/), subject to an application and research proposal meeting the ethical and governance requirements of the Database. For further details on how to apply for access to the data and for a research proposal template please.

## Code availability

Code for model development, analysis and visualisation are available at https://github.com/eyrelab/abx_selection.

