## Supplementary material for "Machine learning to predict antibiotic susceptibility in Enterobacterales bloodstream infections compared to clinician prescribing"

### Machine learning to predict antibiotic susceptibility in blood stream infections compared to clinician prescribing: Supplement

### Supplementary methods

#### Microbiology

Blood cultures were obtained and incubated using the BD BACTEC system (Becton, Dickinson and Company) and antimicrobial susceptibility results determined by broth microdilution using the BD Phoenix platform and EUCAST guidelines.

#### Antimicrobial resistance prediction

We made predictions at two time points, firstly at the time the blood culture was obtained and secondly when the species was identified. As the exact time point that the species was identified was not recorded in our dataset, we conservatively assume that only 24 hours of further data from the point of sampling were available for this prediction.

#### Model architecture and data partitioning

We fit separate models for each antibiotic (both with and without species information as input features). Predictions were undertaken using XGBoost which offers amongst the best performance for similar tasks using structured tabular data of type analysed here. Model fitting was performed using Python 3.12 and SciKitLearn version 1.5.1.

We used a temporal training-test split to mimic real-world implementation. Data from 01-January-2017 to 31-December-2021 were used for model training. Performance was tested using a held-out data from 01-January-2022 to 31-December-2022 (Test dataset 1). Within the training data we used 5-fold cross validation to undertake hyperparameter tuning using Bayesian optimisation (see Table S2 for a list of hyperparameters, ranges explored, and Table S3 for values chosen). We used Platt's method as implemented in SciKitLearn's CalibratedClassifierCV function, to ensure antibiotic resistance probabilities were well calibrated. Thresholds for calling the presence of resistance were chosen within the training data to maximise the sum of sensitivity and specificity.

#### Comparison with clinical decision making

To compare our models with clinical practice, we combined both test datasets and considered patients initially treated with a beta-lactam antibiotic. The antibiotic chosen by clinicians was determined by taking the most recently started beta-lactam with an active prescription within ±4 hours from the time the blood culture was collected. By taking the most recently started beta-lactam we aimed to capture the antibiotic clinicians intended to continue.

Beta-lactams were the most commonly used antibiotics in our institution, and facilitated establishing a hierarchy of antibiotic choices. We included patients empirically treated with amoxicillin, co-amoxiclav, ceftriaxone, piperacillin-tazobactam, or a carbapenem (mostly meropenem; a small number receiving empirical ertapenem), in order of increasing spectrum of coverage. Alternative ordering of ceftriaxone could potentially be considered, but we place it above co-amoxiclav as resistance to ceftriaxone was less common in our setting. After first considering if any active antibiotic (beta-lactam or non-beta-lactam) was given in the baseline period, we then focus on if an active beta-lactam was given. The most common adjunctive antibiotic in our setting was single dose gentamicin, however we exclude it here from our main analysis, as we have previously shown it does not rescue patients with beta-lactam (co-amoxiclav) resistance from associated increases in mortality with *Escherichia coli* bloodstream infection.[1]

We excluded from the clinical comparison neutropenic patients, as from our list of beta-lactams only piperacillin-tazobactam or meropenem would be appropriate empirical therapy. We also exclude patients where no antibiotic was given during the baseline period, as the clinical team was presumed to have taken the blood cultures for diagnostic purposes, but not believed initially there was strong enough evidence to start treatment. Blood cultures also needed to have an available susceptibility result for each of the beta-lactams listed above. No patient allergy data were available.

To compare clinical practice and models predictions, we evaluate the number of patients who are i) optimally treated, i.e. receiving the least broad-spectrum beta-lactam to which their blood culture isolate is sensitive, ii) under-treated, given a beta-lactam with resistance present, and iii) over-treated, given an active beta-lactam, but one that was of a broader spectrum than was necessary. We also describe the relative usage rates of each antibiotic.

We evaluated 4 strategies for applying our machine learning predictions. In strategy 1, we used the training data to identify resistance probability threshold values for each antibiotic that matched the relative use of each antibiotic by the model to use by clinicians. Having set the probability thresholds using the training data, we then applied them to the test data. In strategy 2, we set the prediction thresholds to match the proportion of patients receiving each antibiotic to rates of susceptibility. Because we do not expect our models to perform perfectly, we also perform a sensitivity analysis where we allow a 20% reduction in use of each the two narrowest spectrum agents, amoxicillin and co-amoxiclav, assigning the remaining use proportionally. In strategy 3, we match the number of patients over-treated by clinicians and the algorithm, but otherwise do not constrain antibiotic choices, albeit still favouring the narrowest spectrum agent where possible. Within this constraint we assess if the number of patients receiving active treatment can be increased. As co-amoxiclav is the most commonly used antibiotic, but also has relatively high resistant rates, we also evaluate the simple comparator of assuming the first-line antibiotic guideline was switched to ceftriaxone, such that all patients who received amoxicillin or co-amoxiclav are reassigned to ceftriaxone. To provide a model-based comparison with this approach, in strategy 4 we choose thresholds for predicting resistance, such that we match the overall proportion of patients receiving active treatment, to investigate if over-treatment is reduced by the model.

#### Supplementary tables

| | |
|---|---|
| Antibiotics | For amoxicillin, cefalexin, ceftriaxone, ciprofloxacin, co-amoxiclav, co-trimoxazole, ertapenem, gentamicin, meropenem, nitrofurantoin, piperacillin-tazobactam, and trimethoprim:<br>▪ Number of hospital prescribed courses in last year<br>▪ Time since last hospital prescribed course in last year<br><br>For any antibiotic:<br>▪ Number of hospital prescribed courses in last year<br>▪ Time since last hospital prescribed course in last year |
| Clinical syndrome | Clinical syndrome (derived from antibiotic indication field, binary presence / absence of each): abdominal, ear nose and throat, neurological, no specific source, non-informative text, orthopaedic, other specific, prophylaxis, respiratory, skin and soft tissue, uncertain, urinary |
| Population antibiotic resistance rate | In blood cultures, based on samples in the last year: amoxicillin, ceftriaxone, ciprofloxacin, co-amoxiclav, co-trimoxazole, ertapenem, gentamicin, meropenem, and piperacillin-tazobactam<br><br>Any sample, based on samples in the last year: amoxicillin, ceftriaxone, ciprofloxacin, co-amoxiclav, co-trimoxazole, ertapenem, gentamicin, meropenem, and piperacillin-tazobactam |
| For species analysis only, Species identified | *Citrobacter spp*, *Enterobacter spp*, *Escherichia coli*, *Klebsiella spp*, *Proteus Providencia Morganella spp*, *Serratia spp,* or Other |
| Comorbidity | Age adjusted Charlson score<br>Charlson score<br>Each of the individual domains of the Charlson score (binary presence / absence) |
| Demographics | Age<br>Sex<br>Index of multiple deprivation score (higher=more deprived) |
| Personal history of AMR infections | CRE in last year (binary)<br>ESBL in last year<br>Time since last ESBL in the last year<br>MRSA in last year<br>VRE in last year |
| Index date | Hour of day blood culture taken (as proxy for acuity) |
| Labs | Count of measurements in last 72 hours:<br>▪ Full blood count<br>▪ Renal function<br>▪ Liver function<br>▪ CRP<br>▪ Clotting<br>▪ Blood gas |
| Personal microbiology results | For amoxicillin, cefalexin, ceftriaxone, ciprofloxacin, co-amoxiclav, co-trimoxazole, ertapenem, gentamicin, meropenem, nitrofurantoin, piperacillin-tazobactam, and trimethoprim:<br>▪ Count of isolates from any sample and any species with resistance in last year<br>▪ Time since last resistant isolate of any species in the last year<br><br>Additionally, for the species analysis for the same antibiotics:<br>▪ Count of isolates from any sample of the same species with resistance in last year<br>▪ Time since last resistant isolate of the same species in the last year<br><br>Count of positive blood cultures in last year<br>Count of positive urine cultures in last year |

| | |
|---|---|
| | Time since last positive blood culture in last year<br>Time since last positive urine culture in last year<br><br>Count of blood cultures containing Enterobacterales in last year<br>Count of urine cultures containing Enterobacterales in last year<br>Time since last blood culture with Enterobacterales in the last year<br>Time since last urine culture with Enterobacterales in the last year<br><br>Time since last blood culture sent<br>Time since last urine culture sent |
| Personal factors | Body mass index<br>Height<br>Weight |
| Hospital exposure | Days since start of hospital admission/attendance at blood culture sampling<br><br>Admission type: Elective admission, Emergency admission, Maternity admission, Other admission, Outpatient attendance<br><br>Count of admissions in last 30 days, 90 days and last year<br>Days in hospital in last 30 days, 90 days and last year |
| Procedures | Any clean surgery in last year<br>Any clean contaminated surgery in last year<br>Any contaminated surgery in last year<br>Any urinary catheter code in last year |
| Specialty | Specialty at time of blood culture sampling: Acute and general surgery, Acute, emergency and geriatric medicine, Critical care, Medical subspecialty, Obstetrics, Others, Paediatrics, Surgical subspecialty, Trauma and orthopaedics |
| Vitals | Count of measurements obtained in last 24 hours |

**Table S1. List of model features**. For counts of laboratory haematology and biochemistry tests and vital signs we allowed the window searched to extend back 72 hours and 24 hours respectively, and forward 4 hours to account for results obtained around the same time as blood cultures. In the species analysis we extended this look forward by a further 24 hours, i.e. 28 hours in total to allow for additional tests conducted between blood culture sampling and a species being identified (the exact time of species identification was not available in our dataset).

| Parameter | Range for antibiotics with more events: amoxicillin, co-amoxiclav, co-trimoxazole | Range for antibiotics with fewer events: ceftriaxone, piperacillin-tazobactam, ciprofloxacin, gentamicin |
|---|---|---|
| n_estimators | range(50, 1500, 20) | range(50, 800, 20) |
| learning_rate | [0.0001, 0.0002, 0.0005, 0.001, 0.002, 0.005] | [0.0001, 0.0002, 0.0005, 0.001, 0.002, 0.005] |
| max_depth | range(3,12,1) | range(3,6,1) |
| gamma | range(1,20,1) | range(3,20,1) |
| min_child_weight | range(2,20,1) | range(3,20,1) |
| colsample_bytree | [i/20.0 for i in range(1,16)] | [i/20.0 for i in range(1,16)] |
| subsample | [i/20.0 for i in range(1,16)] | [i/20.0 for i in range(1,16)] |

**Table S2. Hyperparameter search spaces for Bayesian hyperparameter optimisation**. For each antibiotic up to 100 iterations of hyperparameter optimisation were performed. Search spaces are given using python code.

| Antibiotic | Species data included | n_ estimators | learning _rate | max_ depth | gamma | min_ child_ weight | colsample _bytree | subsample |
|---|---|---|---|---|---|---|---|---|
| Amoxicillin | No | 390 | 0.005 | 5 | 7 | 3 | 0.6 | 0.75 |
| Co-amoxiclav | | 690 | 0.002 | 5 | 7 | 2 | 0.35 | 0.75 |
| Ceftriaxone | | 770 | 0.001 | 5 | 9 | 5 | 0.75 | 0.4 |
| Piperacillin-tazobactam | | 750 | 0.0002 | 4 | 3 | 3 | 0.5 | 0.65 |
| Ciprofloxacin | | 670 | 0.005 | 3 | 7 | 10 | 0.55 | 0.6 |
| Co-trimoxazole | | 1090 | 0.002 | 4 | 7 | 6 | 0.45 | 0.7 |
| Gentamicin | | 250 | 0.005 | 3 | 4 | 3 | 0.7 | 0.35 |
| Amoxicillin | Yes | 1230 | 0.002 | 5 | 11 | 4 | 0.75 | 0.75 |
| Co-amoxiclav | | 730 | 0.005 | 4 | 1 | 6 | 0.45 | 0.6 |
| Ceftriaxone | | 750 | 0.002 | 4 | 3 | 3 | 0.45 | 0.6 |
| Piperacillin-tazobactam | | 550 | 0.005 | 3 | 3 | 7 | 0.75 | 0.6 |
| Ciprofloxacin | | 690 | 0.002 | 5 | 3 | 7 | 0.35 | 0.6 |
| Co-trimoxazole | | 1210 | 0.005 | 11 | 6 | 4 | 0.25 | 0.6 |
| Gentamicin | | 750 | 0.005 | 5 | 5 | 7 | 0.65 | 0.7 |

**Table S3. Selected model hyperparameters**.

| Antibiotic | n | Resistant, n | Resistant, % | AUC (95%CI) | Sensitivity (95% CI) | Specificity (95% CI) | Positive predictive value (95% CI) | Negative predictive value (95% CI) |
|---|---|---|---|---|---|---|---|---|
| Amoxicillin | 3260 | 2193 | 67 | 0.739 (0.721 - 0.756) | 62.4 (60.3 - 64.5) | 75.0 (72.2 - 77.5) | 83.7 (81.9 - 85.3) | 49.3 (46.7 - 51.8) |
| Co-amoxiclav | 3257 | 1377 | 42 | 0.751 (0.734 - 0.767) | 63.4 (61.0 - 66.0) | 74.9 (73.0 - 76.8) | 64.9 (62.3 - 67.4) | 73.6 (71.8 - 75.5) |
| Ceftriaxone | 3264 | 363 | 11 | 0.916 (0.901 - 0.929) | 86.5 (82.9 - 89.9) | 80.1 (78.7 - 81.5) | 35.2 (32.2 - 38.6) | 97.9 (97.4 - 98.4) |
| Piperacillin-tazobactam | 3273 | 214 | 7 | 0.944 (0.929 - 0.957) | 88.3 (84.3 - 92.4) | 87.5 (86.3 - 88.6) | 33.1 (28.8 - 37.2) | 99.1 (98.7 - 99.4) |
| Ciprofloxacin | 3275 | 414 | 13 | 0.908 (0.896 - 0.920) | 86.2 (82.6 - 89.4) | 78.9 (77.5 - 80.5) | 37.2 (34.2 - 40.3) | 97.5 (96.9 - 98.1) |
| Co-trimoxazole | 3202 | 761 | 24 | 0.942 (0.934 - 0.950) | 89.4 (87.0 - 91.5) | 87.8 (86.4 - 89.0) | 69.5 (66.4 - 72.5) | 96.4 (95.5 - 97.1) |
| Gentamicin | 3266 | 307 | 9 | 0.834 (0.812 - 0.858) | 81.8 (77.2 - 86.1) | 71.1 (69.5 - 72.8) | 22.7 (20.4 - 25.4) | 97.4 (96.7 - 98.1) |

**Table S4. Model performance for predicting antibiotic resistance at blood culture sampling in training dataset 1, 01 January 2017 – 31 December 2021**. AUC, area under the receiver operating curve. Confidence intervals were generated by bootstrapping with 1000 iterations.

| Antibiotic | n | Resistant, n | Resistant, % | AUC (95%CI) | Sensitivity (95% CI) | Specificity (95% CI) | Positive predictive value (95% CI) | Negative predictive value (95% CI) |
|---|---|---|---|---|---|---|---|---|
| Amoxicillin | 3260 | 2193 | 67 | 0.680 (0.639 - 0.721) | 80.9 (77.3 - 84.5) | 42.4 (36.2 - 48.4) | 72.9 (69.1 - 76.5) | 53.7 (47.1 - 60.9) |
| Co-amoxiclav | 3257 | 1377 | 42 | 0.684 (0.643 - 0.725) | 79.5 (74.7 - 83.9) | 39.2 (34.7 - 43.8) | 47.1 (42.6 - 51.4) | 73.8 (67.8 - 79.5) |
| Ceftriaxone | 3264 | 363 | 11 | 0.737 (0.672 - 0.798) | 79.5 (69.9 - 88.7) | 45.7 (41.8 - 49.7) | 15.5 (11.9 - 19.3) | 94.7 (92.2 - 97.2) |
| Piperacillin-tazobactam | 3273 | 214 | 7 | 0.708 (0.641 - 0.781) | 85.9 (77.6 - 94.0) | 37.7 (33.9 - 41.6) | 12.1 (9.1 - 15.1) | 96.4 (93.9 - 98.5) |
| Ciprofloxacin | 3275 | 414 | 13 | 0.726 (0.657 - 0.790) | 76.7 (67.4 - 85.4) | 44.0 (40.1 - 48.1) | 16.0 (12.8 - 19.5) | 93.2 (90.2 - 95.9) |
| Co-trimoxazole | 3202 | 761 | 24 | 0.698 (0.636 - 0.756) | 70.7 (61.8 - 78.9) | 52.6 (48.6 - 56.5) | 24.5 (20.1 - 28.9) | 89.2 (85.6 - 92.7) |
| Gentamicin | 3266 | 307 | 9 | 0.700 (0.628 - 0.775) | 72.0 (62.0 - 82.5) | 51.2 (47.3 - 55.1) | 15.0 (11.5 - 18.7) | 93.9 (91.2 - 96.3) |

**Table S5. Model performance for predicting antibiotic resistance at blood culture sampling in held-out test dataset 1, 01 January 2022 – 31 December 2022 targeting 80% sensitivity**. AUC, area under the receiver operating curve. Confidence intervals were generated by bootstrapping with 1000 iterations. Thresholds for determining resistance set to target sensitivity of 80% using test dataset 2 (used as a separate validation dataset for this purpose).

| Antibiotic | n | Resistant, n | Resistant, % | AUC (95%CI) with species information | Sensitivity (95% CI) | Specificity (95% CI) | Positive predictive value (95% CI) | Negative predictive value (95% CI) |
|---|---|---|---|---|---|---|---|---|
| Amoxicillin | 3260 | 2193 | 67 | 0.823 (0.810 - 0.836) | 62.7 (60.7 - 64.7) | 89.8 (87.8 - 91.5) | 92.7 (91.1 - 93.9) | 54.0 (51.6 - 56.3) |
| Co-amoxiclav | 3257 | 1377 | 42 | 0.819 (0.805 - 0.834) | 68.0 (65.7 - 70.5) | 80.1 (78.4 - 81.9) | 71.5 (69.0 - 74.0) | 77.4 (75.6 - 79.3) |
| Ceftriaxone | 3264 | 363 | 11 | 0.914 (0.898 - 0.928) | 90.1 (87.0 - 93.3) | 74.5 (72.9 - 76.1) | 30.6 (28.0 - 33.5) | 98.4 (97.8 - 98.9) |
| Piperacillin-tazobactam | 3273 | 214 | 7 | 0.901 (0.879 - 0.923) | 86.4 (81.9 - 91.1) | 76.9 (75.4 - 78.4) | 20.7 (18.1 - 23.4) | 98.8 (98.3 - 99.2) |
| Ciprofloxacin | 3275 | 414 | 13 | 0.911 (0.897 - 0.923) | 85.3 (81.6 - 88.6) | 80.0 (78.6 - 81.4) | 38.1 (35.0 - 41.3) | 97.4 (96.7 - 98.0) |
| Co-trimoxazole | 3202 | 761 | 24 | 0.927 (0.917 - 0.936) | 86.7 (84.3 - 89.1) | 84.3 (82.8 - 85.7) | 63.3 (60.4 - 66.4) | 95.3 (94.4 - 96.2) |
| Gentamicin | 3266 | 307 | 9 | 0.883 (0.864 - 0.903) | 75.2 (70.3 - 80.2) | 84.8 (83.5 - 86.0) | 33.9 (30.4 - 37.5) | 97.1 (96.4 - 97.7) |

**Table S6. Model performance for predicting antibiotic resistance at blood culture species identification in training dataset 1, 01 January 2017 – 31 December 2021**. AUC, area under the receiver operating curve. Confidence intervals were generated by bootstrapping with 1000 iterations.

#### Supplementary figures

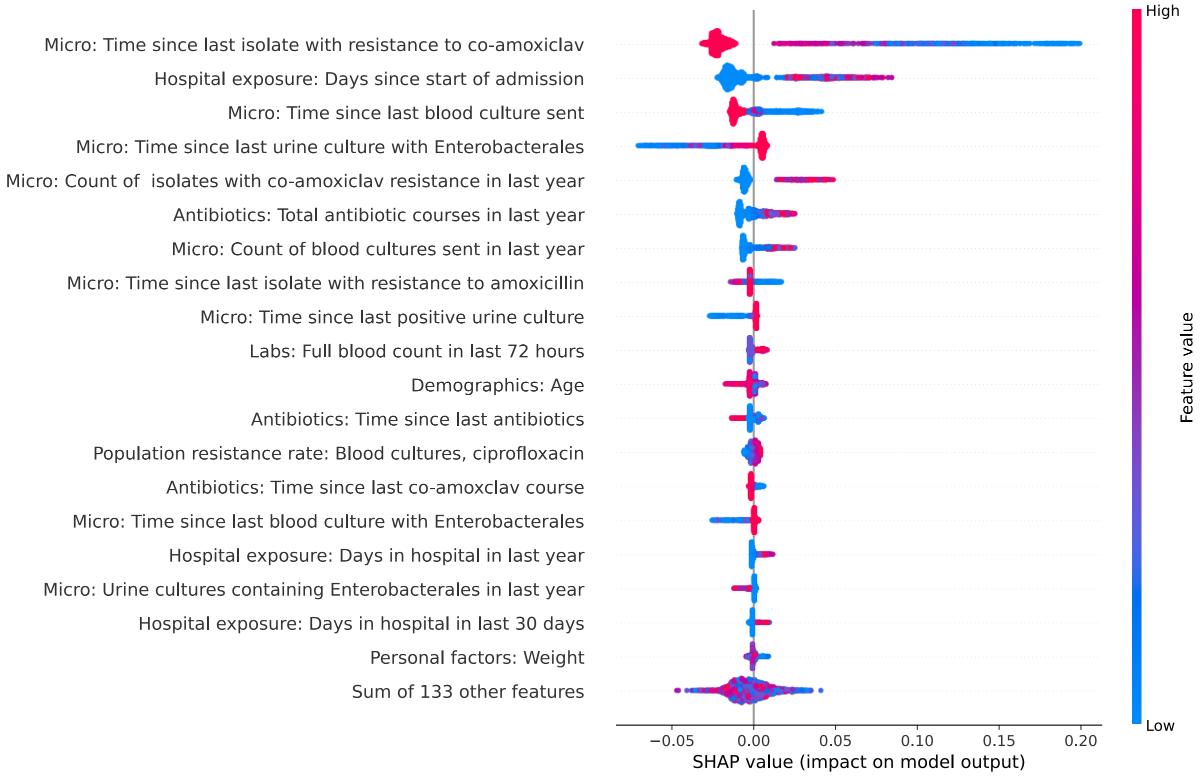

**Figure S1. SHAP (SHapley Additive exPlanations) plot showing feature importance and impacts on model output for predicting co-amoxiclav resistance at blood culture sampling.**

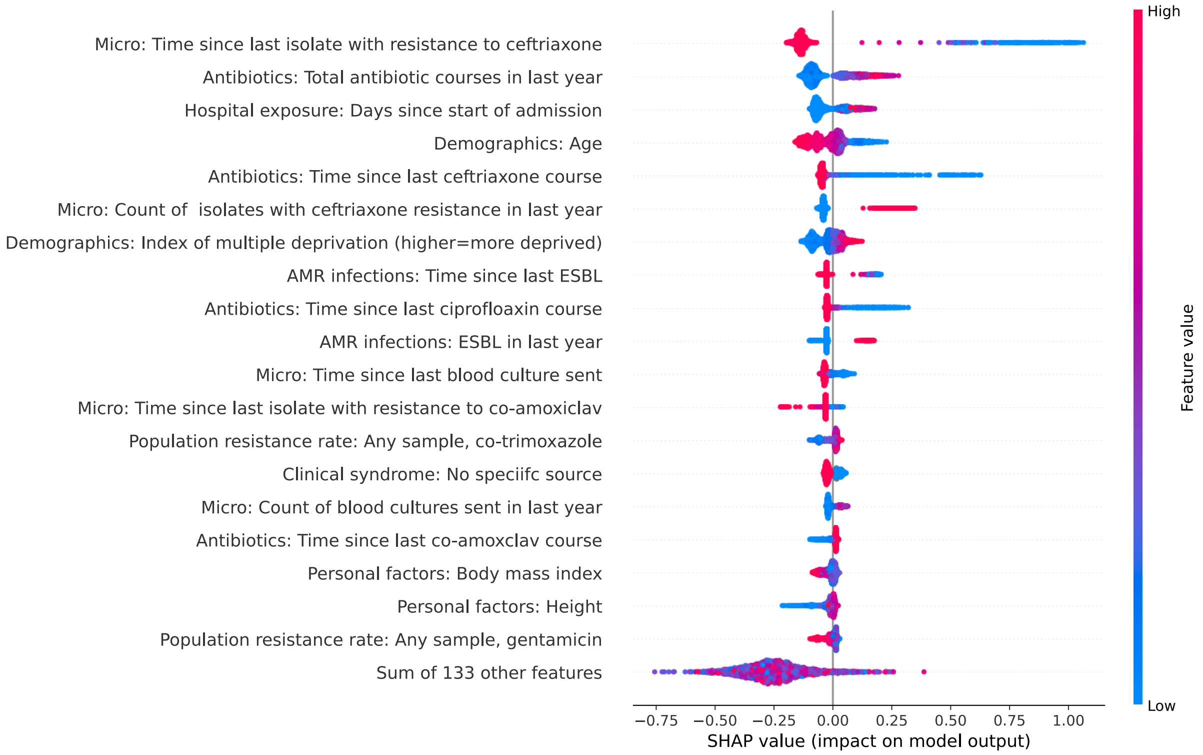

**Figure S2. SHAP (SHapley Additive exPlanations) plot showing feature importance and impacts on model output for predicting ceftriaxone resistance at blood culture sampling.**

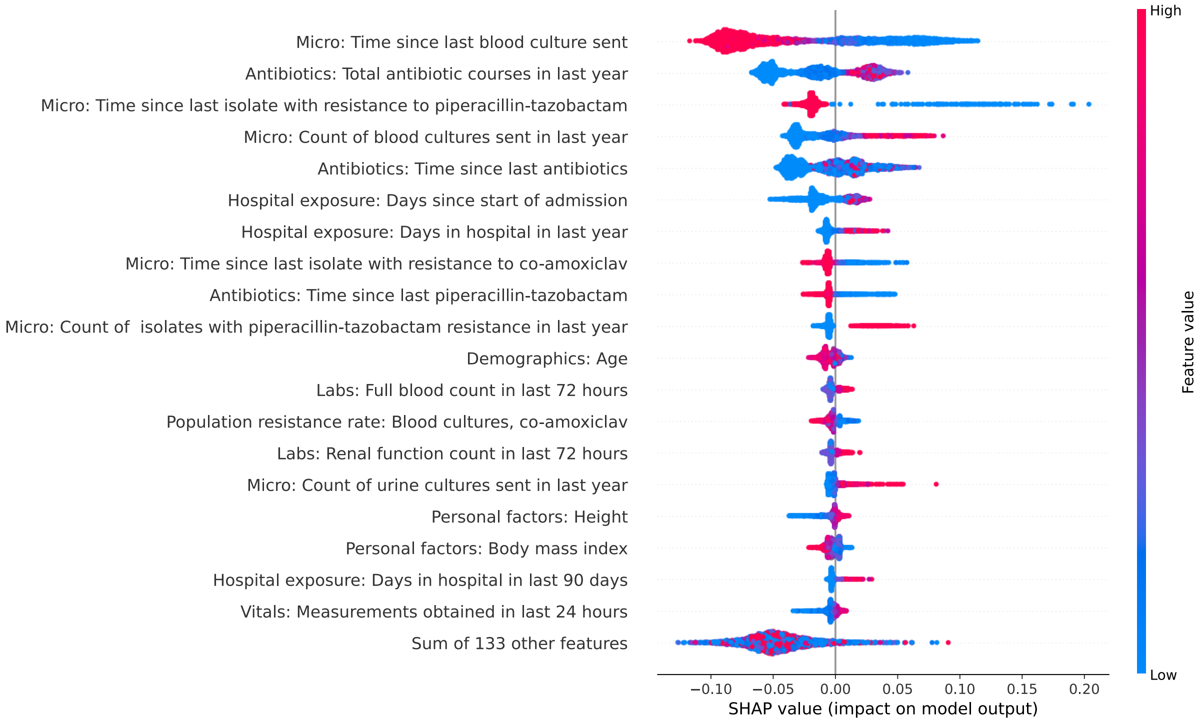

**Figure S3. SHAP (SHapley Additive exPlanations) plot showing feature importance and impacts on model output for predicting piperacillin-tazobactam resistance at blood culture sampling.**

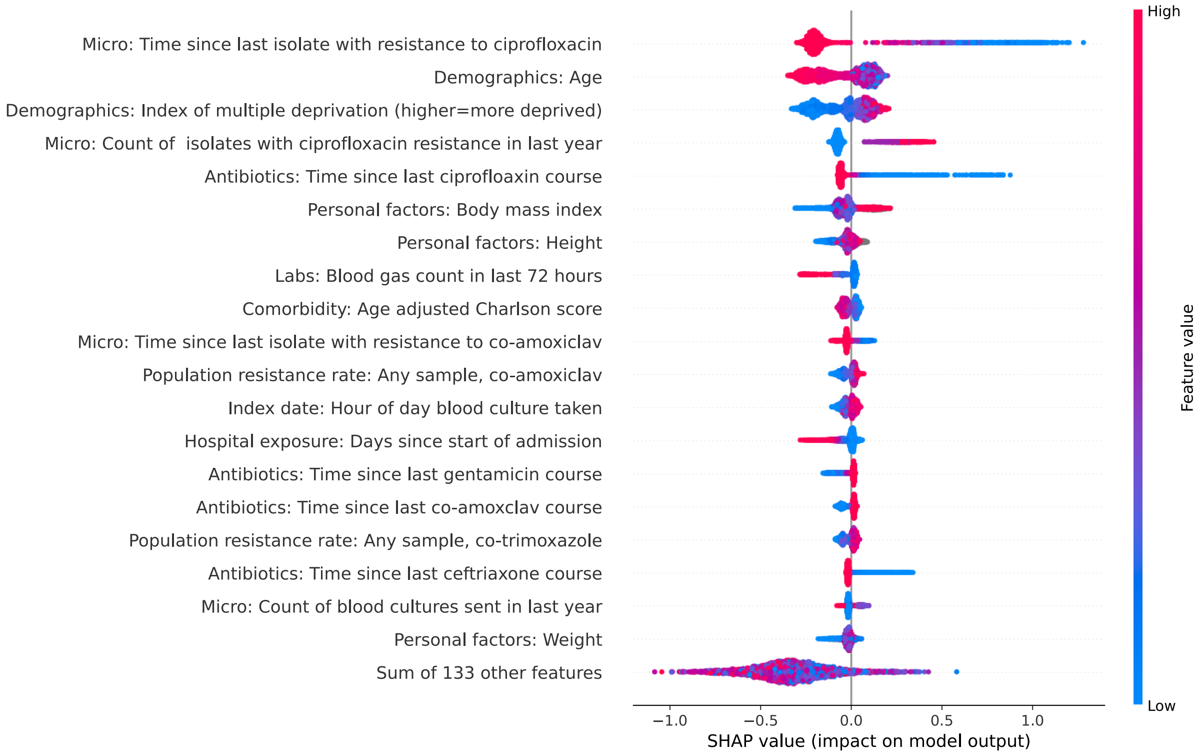

**Figure S4. SHAP (SHapley Additive exPlanations) plot showing feature importance and impacts on model output for predicting ciprofloxacin resistance at blood culture sampling.**

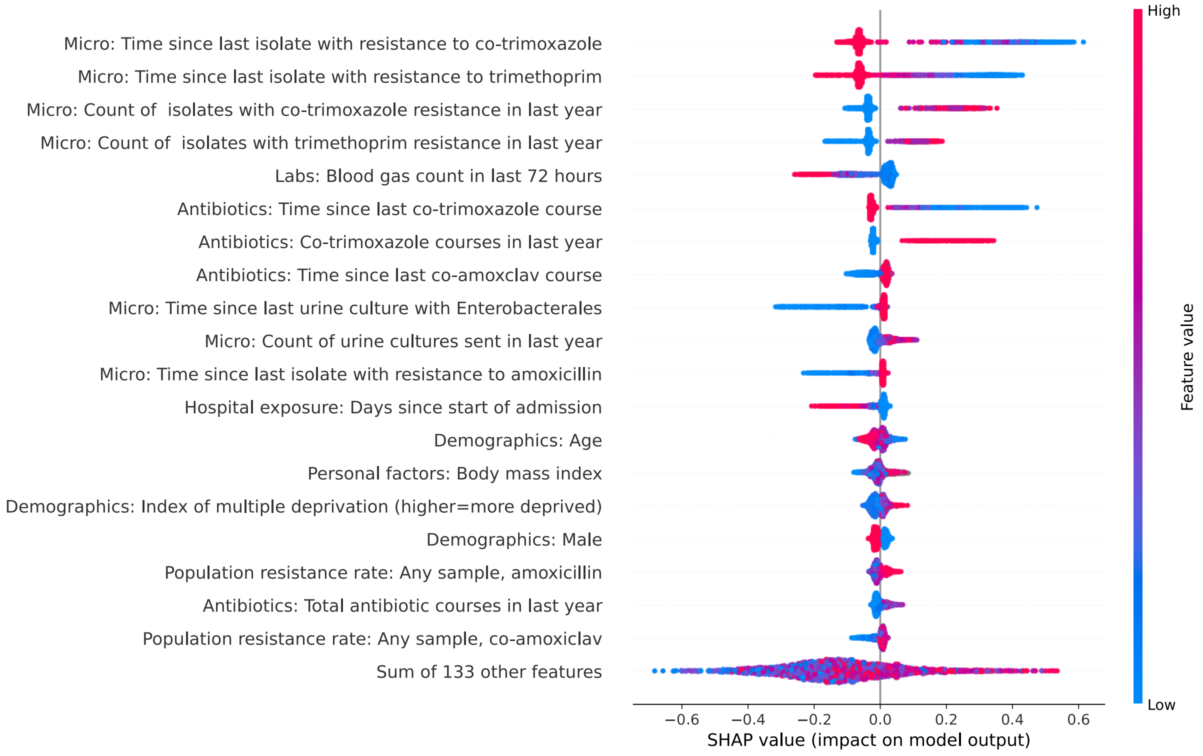

**Figure S5. SHAP (SHapley Additive exPlanations) plot showing feature importance and impacts on model output for predicting co-trimoxazole resistance at blood culture sampling.**

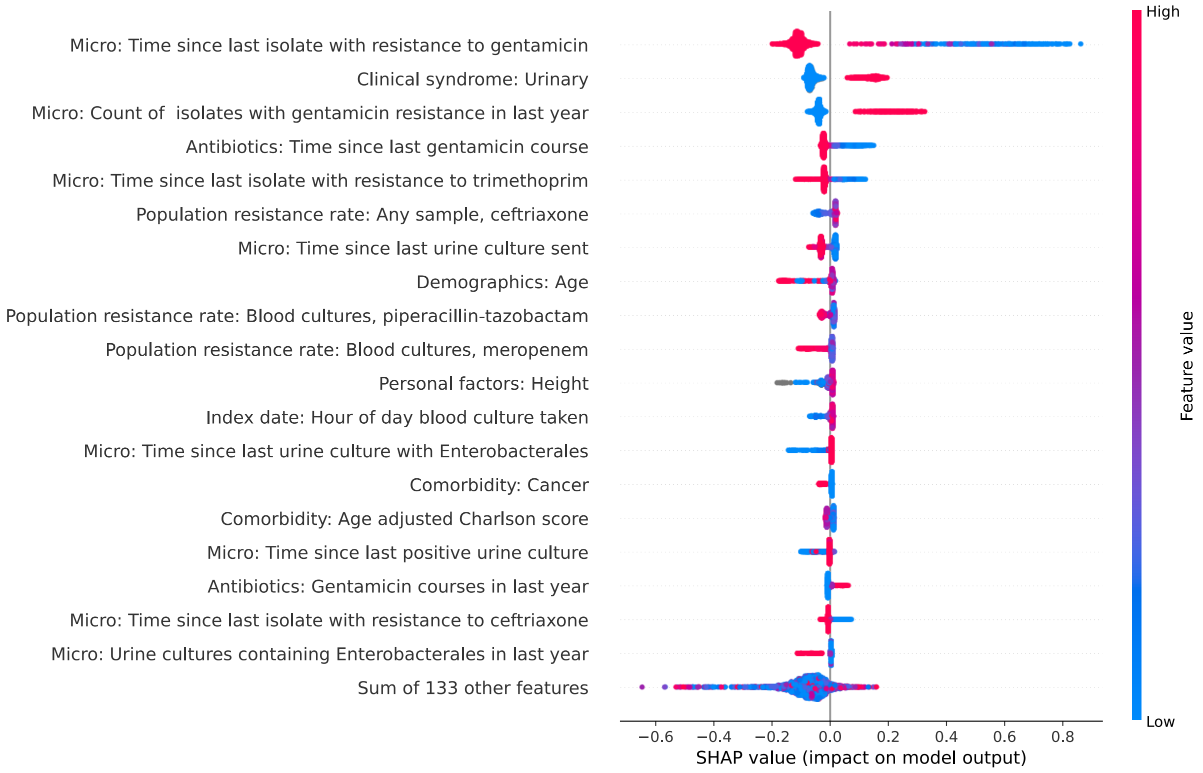

**Figure S6. SHAP (SHapley Additive exPlanations) plot showing feature importance and impacts on model output for predicting gentamicin resistance at blood culture sampling.**

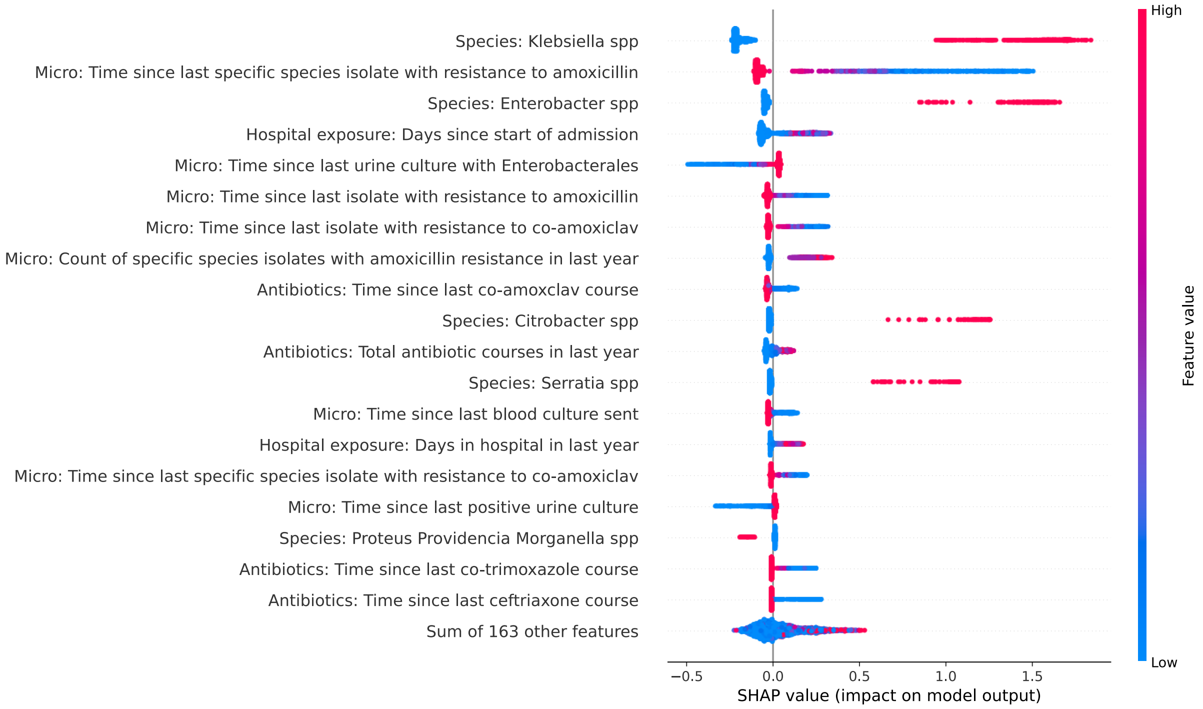

**Figure S7. SHAP (SHapley Additive exPlanations) plot showing feature importance and impacts on model output for predicting amoxicillin resistance at blood culture species identification.**

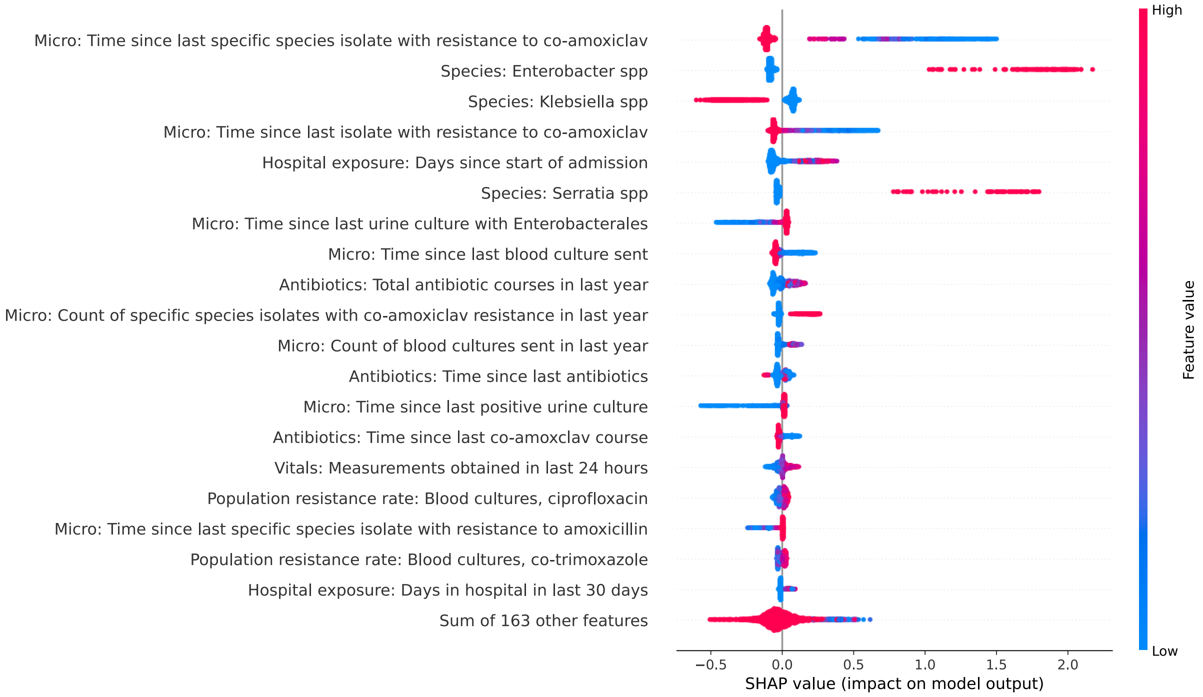

**Figure S8. SHAP (SHapley Additive exPlanations) plot showing feature importance and impacts on model output for predicting co-amoxiclav resistance at blood culture species identification.**

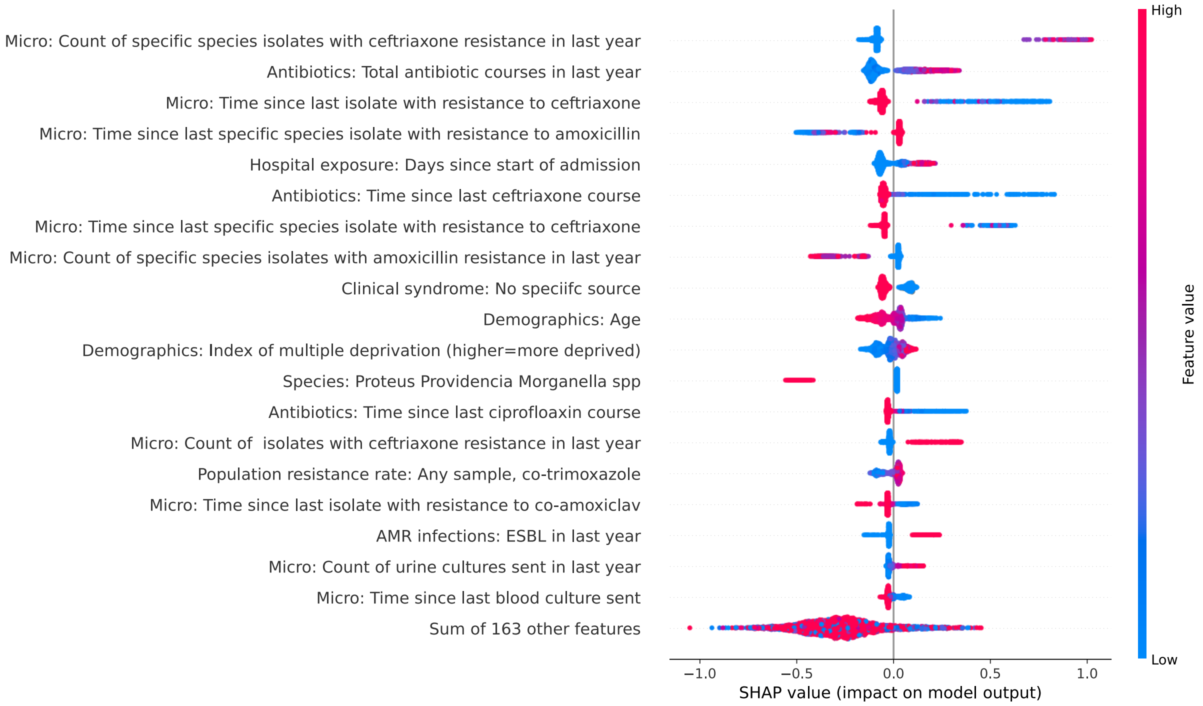

**Figure S9. SHAP (SHapley Additive exPlanations) plot showing feature importance and impacts on model output for predicting ceftriaxone resistance at blood culture species identification.**

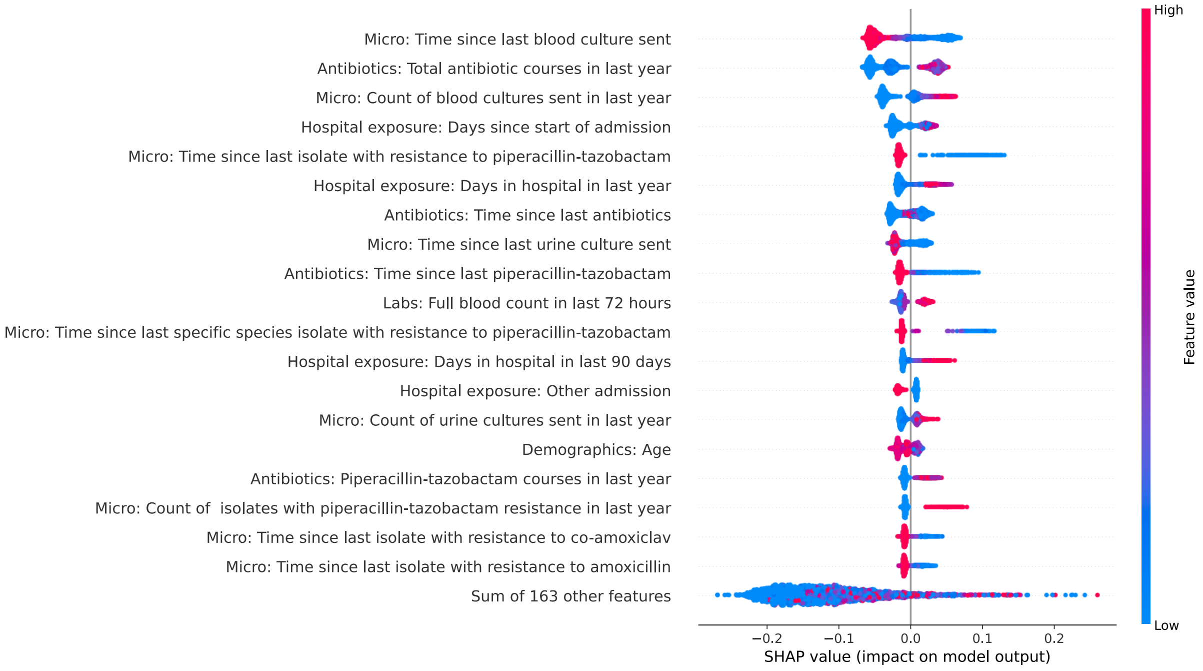

**Figure S10. SHAP (SHapley Additive exPlanations) plot showing feature importance and impacts on model output for predicting piperacillin-tazobactam resistance at blood culture species identification.**

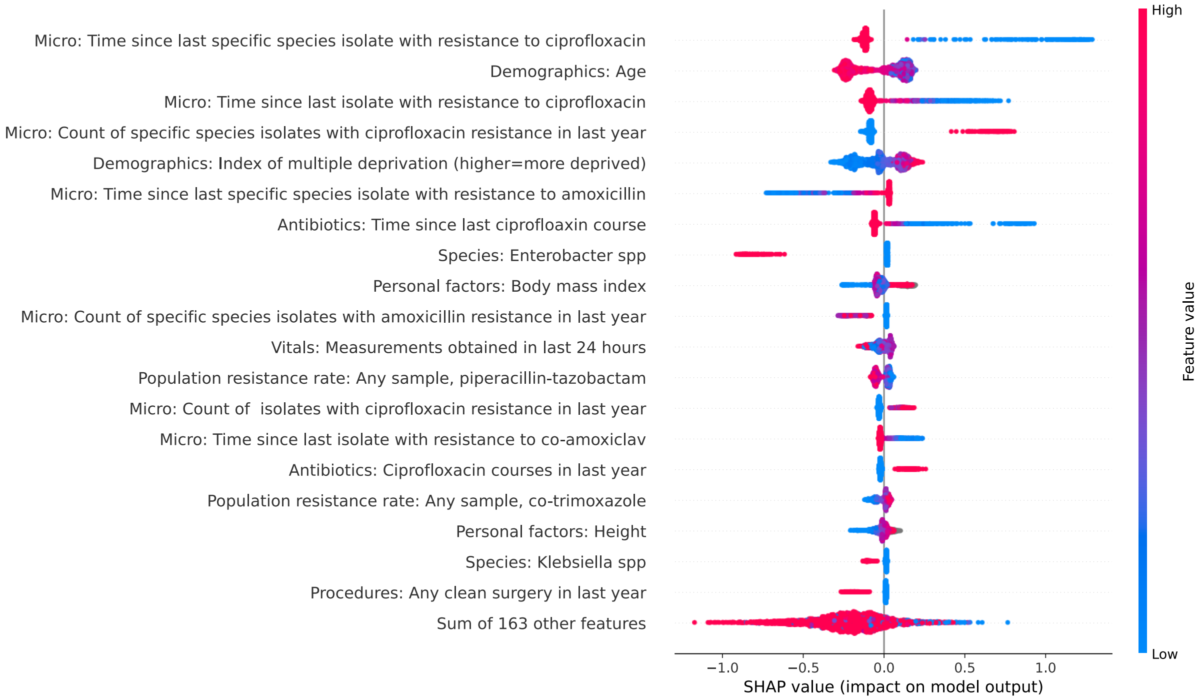

**Figure S11. SHAP (SHapley Additive exPlanations) plot showing feature importance and impacts on model output for predicting ciprofloxacin resistance at blood culture species identification.**

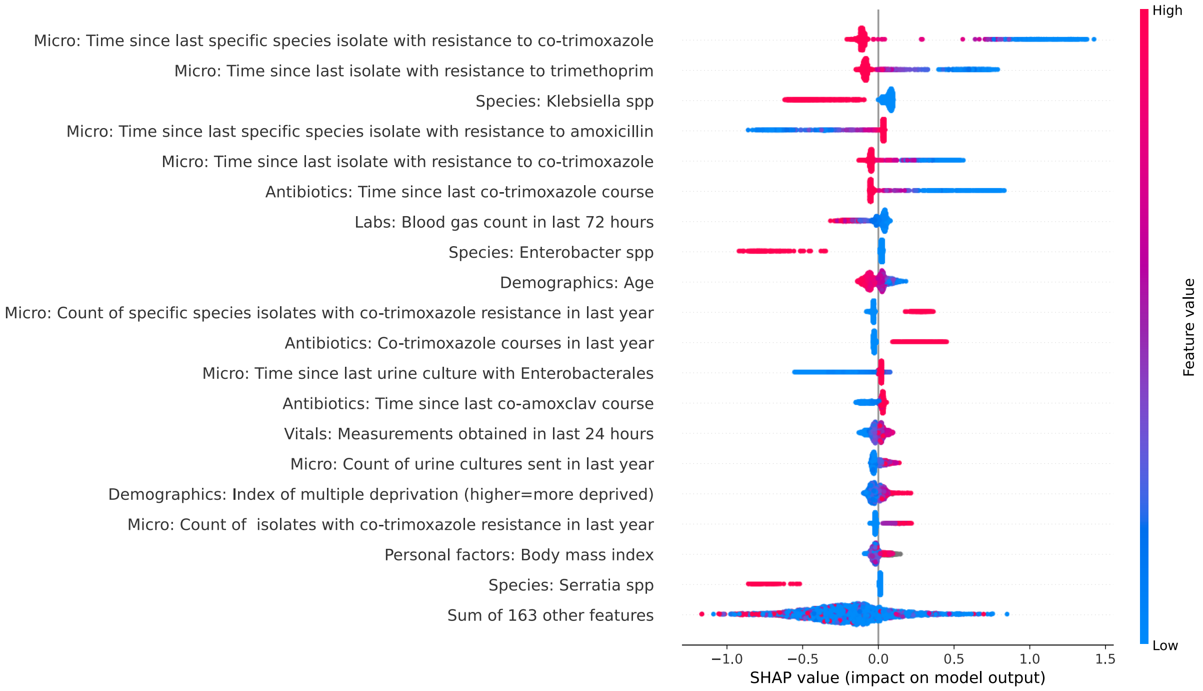

**Figure S12. SHAP (SHapley Additive exPlanations) plot showing feature importance and impacts on model output for predicting co-trimoxazole resistance at blood culture species identification.**

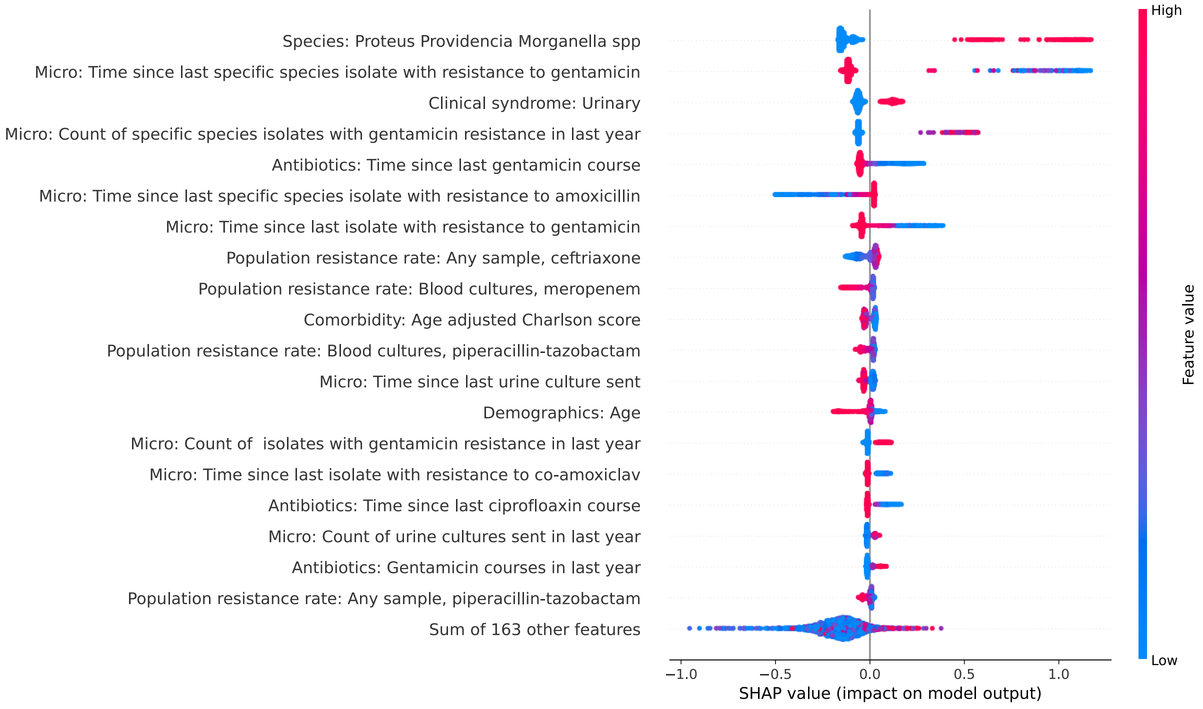

**Figure S13. SHAP (SHapley Additive exPlanations) plot showing feature importance and impacts on model output for predicting gentamicin resistance at blood culture species identification.**

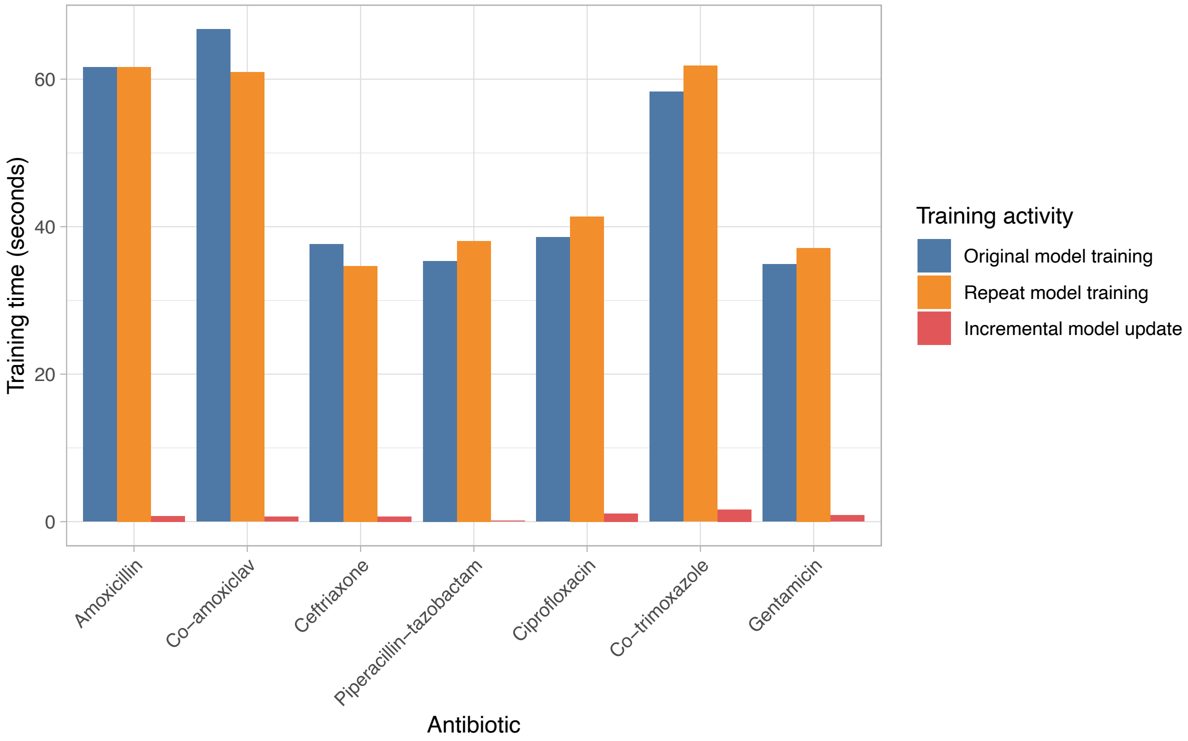

**Figure S14. Model training, retraining and model updating times.** All times are from training using a single Apple M3 Max core. Original and repeat training include hyperparameter optimisation, incremental updates are based on previously obtained hyperparameters from initial training.
